# Fine resolution clustering of *TP53* variants into functional classes predicts cancer risks and spectra among germline variant carriers

**DOI:** 10.1101/2023.12.23.23300440

**Authors:** Emilie Montellier, Nathanaël Lemonnier, Judith Penkert, Claire Freycon, Sandrine Blanchet, Amina Amadou, Florent Chuffart, Nicholas Fischer, Maria Isabel Achatz, Arnold Levine, Catherine Goudie, David Malkin, Gaëlle Bougeard, Christian Kratz, Pierre Hainaut

## Abstract

Li-Fraumeni syndrome (LFS) is a heterogeneous predisposition to a broad spectrum of cancers caused by pathogenic *TP53* germline variants. We have used a clustering approach to assign missense variants to functional classes with distinct quantitative and qualitative features based on transcriptional activity in yeast assays. Genotype-phenotype correlations were analyzed using the germline *TP53* mutation database (n= 3,446) and validated in three LFS clinical cohorts (n= 821). Carriers of class A variants recapitulated all traits of fully penetrant LFS (median age at first diagnosis = 28 years). Class B carriers showed a less penetrant form (median = 33 years, p < 0.05) dominated by adrenocortical and breast cancers. Class C or D carriers had attenuated phenotypes (median = 41 years, p < 0.001) with typical LFS cancers in C and mostly non-LFS cancers in D. This new classification provides insight into structural/functional features causing pathogenicity.

## INTRODUCTION

Li-Fraumeni syndrome (LFS; Mendelian Inheritance in Man [MIM] #151623) is an autosomal dominant cancer predisposition syndrome associated with a high lifetime risk of a broad spectrum of cancers caused by pathogenic or likely pathogenic (P/LP) *TP53* germline (or mosaic) variants, 80% of which are missense ^1^. Typically, LFS occurs in three phases, (1) the childhood phase (0-18 years), characterized by a quartet of early life cancers (adrenocortical carcinoma, ACC; choroid plexus tumor, CPT; medulloblastoma, MB, and rhabdomyosarcoma, RMS, all of which occur almost exclusively before age ten years), and a high risk of soft tissue sarcoma (STS), osteosarcoma (OS), and central nervous system tumors (CNS) in adolescence; (2) the early adult phase, characterized by premenopausal breast cancer (PBC), multiple STS and CNS; and (3) the late adult phase, characterized by lung adenocarcinoma (LUAD), STS, colorectal cancer and prostate cancer ^2^. Clinical definitions are traditionally based on classic LFS criteria ^3^, whereas revised Chompret criteria ^4^ are aimed at identifying patients for *TP53* mutation testing, covering both childhood and adult common presentations of LFS. A definition of the phenotypic spectrum of LFS has been proposed, encompassing (1) phenotypic LFS, defined by the absence of a germline *TP53* variant in persons/families meeting clinical LFS criteria, i.e., classic LFS or Chompret Category A testing criteria; (2) LFS, defined by the presence of a *TP53* variant in persons/families meeting LFS testing criteria or with cancer < 18 years; (3) attenuated LFS (aLFS), defined by the presence of a *TP53* variant in a person/family with cancer ≥ 18 years who does not meet LFS testing criteria; and (4) incidental LFS, defined by the presence of a *TP53* variant in a person/family without a history of cancer ^5^. This phenotypic definition provides a framework to investigate genotype-phenotype correlations by classifying *TP53* variants according to their pathogenicity.

The p53 protein is a multi-functional transcription factor regulating a complex network of cellular and systemic anti-proliferative responses ^6–8^. Loss of these functions, often caused by inactivating missense mutations in the *TP53* gene, impairs several coordinated mechanisms of growth suppression that normally operate to counteract carcinogenesis ^9^. The main functional feature of tumor-associated somatic mutations is the disruption of DNA binding and transactivation capacity of the p53 tumor suppressor protein, either by direct mutation and structural alteration of the DNA-binding domain (DBD), or by destabilization of the oligomerization domain required for high-affinity DNA-binding (Loss of Function, LOF). Stable mutant proteins can also exert dominant-negative effects (DNE) over wild-type allele products and have also been proposed to exert a number of pro-oncogenic gain of function (GOF) effects documented in experimental cell and animal models ^10^, the phenotypic consequences of which remain unclear in the context of LFS. Systematic large-scale studies have assessed the impact of thousands of missense variants on biochemical and biological p53 protein functions in yeast or cell-based experimental assays ^11–13^. This wealth of information, as well as carriers’ phenotypic traits, is used by the ClinGen *TP53* expert panel to inform variant interpretation for clinical purposes (https://clinicalgenome.org/affiliation/50013/) ^14^. Of particular interest among functional datasets is the *TP53* variant transactivation dataset developed over 20 years ago by C. Ishioka et al. using a yeast-based functional assay ^12^. This assay analyzed the transactivation capacity of a panel of 2,314 variants towards synthetic reporters controlled by eight different p53 DNA response elements (p53RE). Thus, this dataset provides the equivalent of eight different “mugshots” for each variant, revealing both quantitative and qualitative features that capture subtle variations in their capacity to bind and transactivate different sequences matching the p53 DNA-binding consensus. In this study, we described and tested a new classification of *TP53* missense variants based on a revisited analysis of Kato’s yeast-based transactivation (YTA) data (**Figure 1, graphical abstract**). We have applied an iterative clustering approach to separate *TP53* missense variants into four classes (A, B, C, D) as compared to nonsense/frameshift variants (class 0) that are considered as completely inactivating the p53 protein. Mapping these classes on phenotypic LFS data from the public repository of germline *TP53* variants curated at NCI (https://tp53.isb-cgc.org/) revealed that each class is associated with defined phenotypic traits within the LFS spectrum. Our results provide a refined resolution of genotype-phenotype correlations in LFS as well as insights on structural/functional features that specify variant pathogenicity.

**Figure 1:**
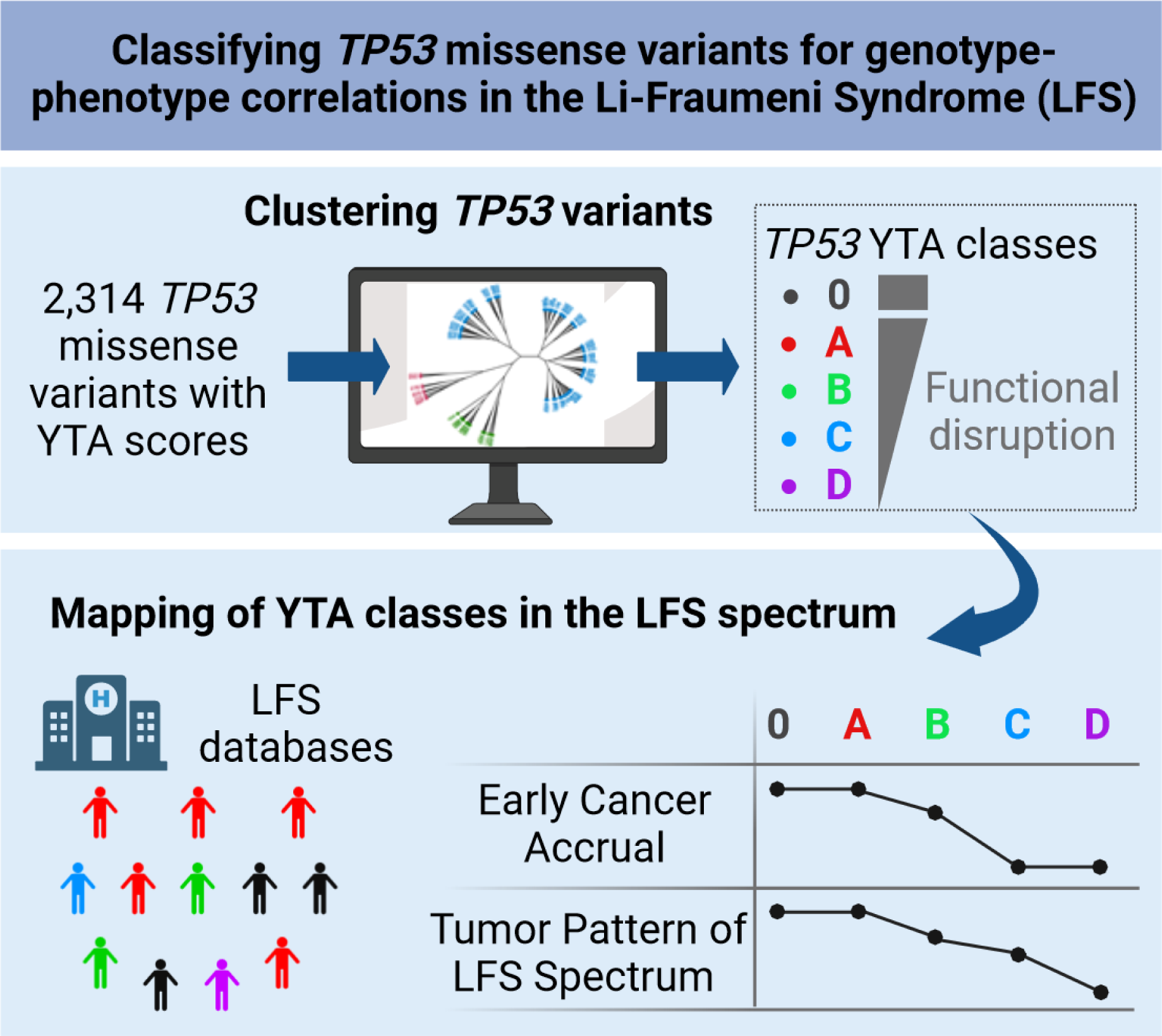
Summary / Graphical Abstract.

## RESULTS & DISCUSSION

### *TP53* variant clustering

An iterative hierarchical Ward’s clustering method was used to interrogate the missense variant dataset developed by Ishioka and collaborators in yeast based functional assays ^12^. We identified 16 iterative clusters of various sizes based on variant similarities in their capacity to differentially activate eight promoters containing different versions of p53 DNA consensus elements (from 613 variants in Cluster 2_2_1 to 5 variants in Cluster 3) (**Figure S1 and Figure S2A**). Each cluster differed by the quantitative and qualitative features of their transcriptional activity, ranging from complete loss of activity towards all promoters (cluster 1_1_1, “triple_1”) to enhanced transcriptional activity (compared to wild-type p53) towards at least some promoters (cluster 3, “supertrans”), with a broad diversity of clusters with intermediate qualitative and quantitative features.

To map these clusters onto the LFS tumor spectrum, we grouped them into four classes (A to D), broadly reflecting a gradient of activity with A having the lowest overall transcriptional activity and D the highest, and B and C displaying intermediate and heterogeneous activities towards different reporters (**Figure S2**). The list of the 2,314 TP53 missense variants and the corresponding clusters and classes is presented in **Table S1**. Classes showed clear differences in variant distribution within the p53 protein structure (**Figure 2A and S3**) and in their predicted structural/functional effects (**Figure S4**). When compared with biophysical prediction scores (SIFT, AGVGD) ^15,16^ and integrative structural and functional scores ^17^, classes highlighted a gradient from A to D, with A and B being enriched in deleterious and non-functional features while C and D were enriched in non-deleterious and functional features. Class A included all common *TP53* cancer mutation hotspots and was enriched in variants at residues located within the surface of the p53 protein in direct contact with DNA, in structural elements supporting the DNA binding surface, or in structural motifs required for the cohesion of the p53 tetramerization domain. Class B included variants mapping to the DBD of p53, albeit at different residues than class A, mostly located within defined sections of the beta-sheets that constitute the scaffold of the DBD as well as in loops exposed at the surface of the protein but not within its DNA-binding surface. Class C and D were enriched in variants that mapped to the N-terminus or to the extreme C-terminus of p53. Class C, in particular, was enriched in variants at residues of the C-terminal regulatory domain with multiple post-translational regulatory sites poorly represented in other classes.

**Figure 2:**
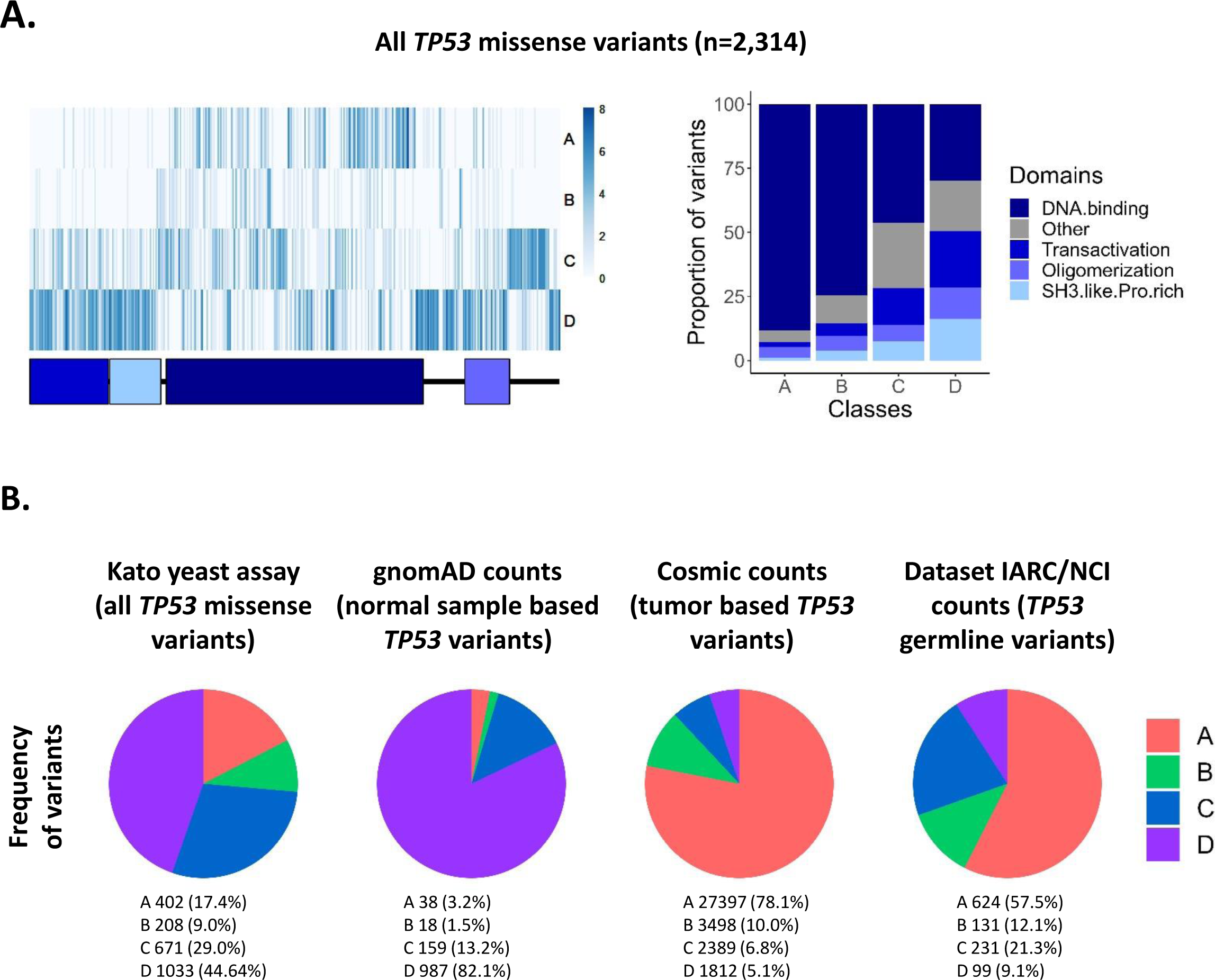
Distribution of *TP53* variants from YTA classes across *TP53* structure and datasets. **A.** Localization of *TP53* missense variants along the *TP53* sequence (left panel). Heatmap showing the number of variants found at each amino acid position, for each YTA classes. The *TP53* domains are indicated below the heatmap to visualize the localization of *TP53* variants within *TP53* secondary structure. Proportion of residues within each *TP53* domain for the four YTA classes (right panel). **B.** Distribution of the variants from the YTA classes within different databases: transactivation yeast assay, gnomAD database, Cosmic database, and IARC/NCI LFS dataset. Pie charts represent the number of samples with *TP53* missense variants belonging to the four YTA classes.

We next mapped the distribution of classes A-D in three *TP53* variant datasets: cancer-related germline variants (NCI; https://tp53.isb-cgc.org/), cancer related somatic variants (COSMIC; https://cancer.sanger.ac.uk/cosmic) and non-cancer related germline variants (gnomAD; https://gnomad.broadinstitute.org/) (**Figure 2B**). Class A variants were enriched by factors of 4.5 and 3.3 fold in the COSMIC and NCI datasets, respectively but were counter-selected (0.2 fold) in the gnomAD dataset. Likewise, albeit to a lesser extent, class B variants showed positive selection in COSMIC (1.1) and NCI (1.3) datasets, but not in gnomAD (0.2). In contrast, class D variants were enriched in gnomAD (1.8) and counter-selected in COSMIC (0.1) and NCI (0.2), whereas class C was not enriched in any of the datasets. These results are compatible with the notion that classes A to D correspond to a gradient of cancer-related p53 dysfunction.

### Variant classes are associated with distinct phenotypic patterns within the LF spectrum

To determine whether variant classes could predict tumor phenotypes within the broad LF spectrum, we analyzed lifetime cancer accrual and tumor patterns using data compiled in the NCI germline *TP53* mutation database (https://tp53.isb-cgc.org/) ^18^. This database assembles information retrieved from the literature on 3,446 individuals (1,522 families) with germline *TP53* variants (4,031 cancer diagnoses). As a basis for comparison, we added a class “zero” (0) that included variants considered as “null” for p53, i.e., non-missense variants (nonsense, frameshift,) that disrupt the production of a functional p53 protein (**Figure S5**). Class 0 thus includes variants considered to have completely lost wild-type p53 function. **Figure 3A** shows that class A variants were found in 1426 (53%) patients, followed by class 0 (552, 21%), B (290, 11%), C (242, 9%), and D (171, 6%). Gender distribution differed among classes, with a lower proportion of males in classes B, C and D (31.4%, 26.7%, and 32%, respectively) than in classes 0 and A (40.9% and 40.6%, respectively) (**Figure 3B**). With respect to lifetime cancer accrual, classes 0 and A were associated with the most severe profiles (median age at first diagnosis = 28 years) (**Figure 3C-E**). Classes C and D were associated with attenuated accrual profiles (median age = 41 years), whereas class B showed an intermediate and distinct profile (median age = 33 years), characterized by rapid accrual during childhood (similar to classes A and 0) and slower accrual during adolescence and adulthood. This gradient of pathogenicity was reflected in the proportion of carriers with multiple cancers as well as cancer free (**Figure 3F-G**) and was consistent with current definitions of clinical phenotypes (**Figure I-J**). Of note, Variants in established cancer predisposition genes other than *TP53* were more frequently found in carriers of class D variants than in carriers of any other variant class (**Figure 3H**).

**Figure 3:**
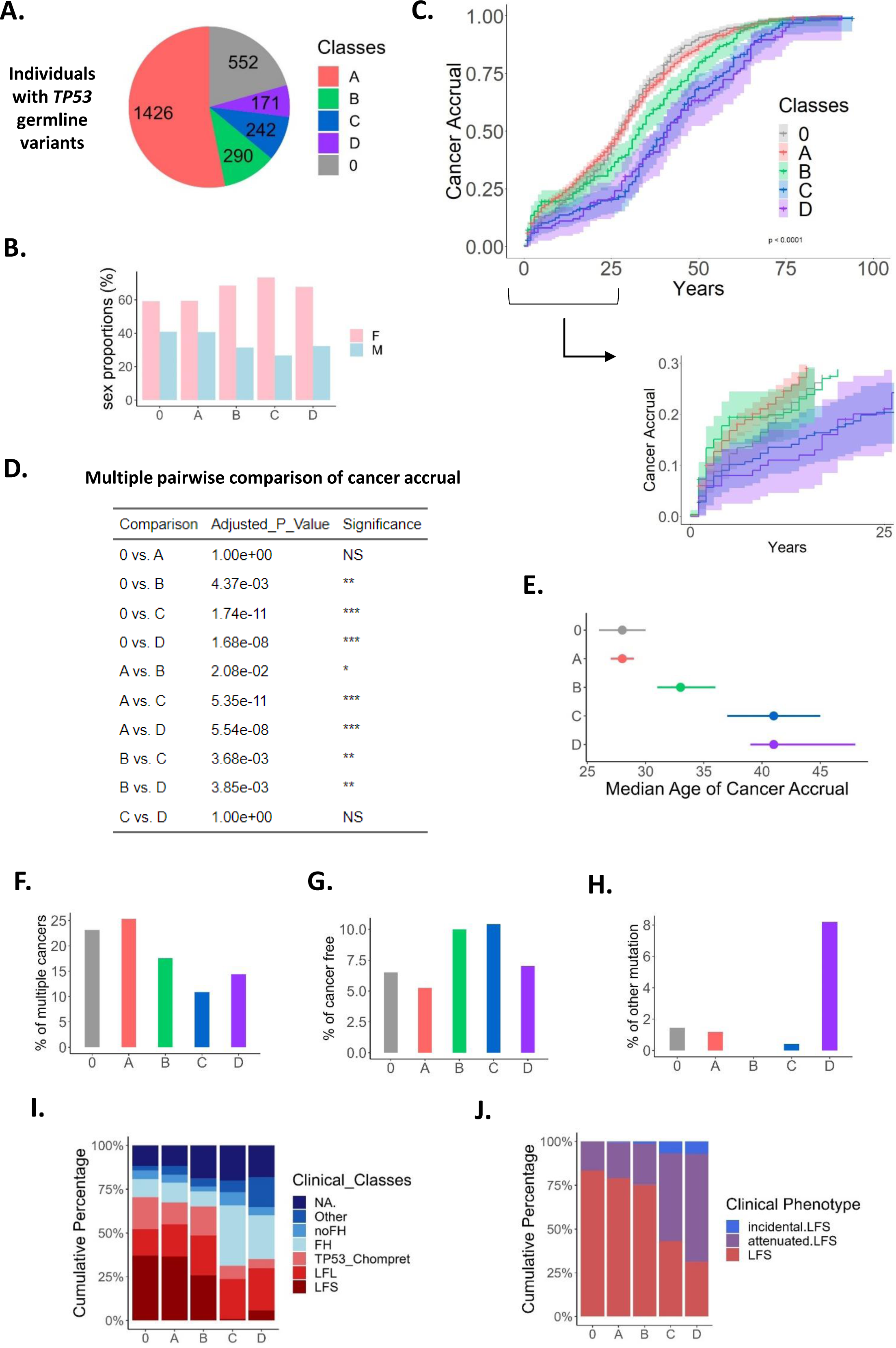
Relationship between YTA classes and clinical phenotype in the Li-Fraumeni Syndrome. **A.** Distribution of *TP53* germline carriers in the IARC/NCI database into classes. Class 0 includes non-missense variants (stop and frameshift). **B.** Sex distribution of individuals (F=females, M=males) for each class. **C.** Cancer accrual of individuals according to classes. The inverted Kaplan-Meier presentation corresponds to the age of onset of the first cancer in each individual. The confidence intervals at 95% are displayed on the curves, and the p-value of the Log-Rank test is indicated. **D.** Pairwise comparison of cancer accrual for each class. A multiple pairwise comparison (with Bonferroni correction) shows the significance of differences in cancer accrual between the classes (adjusted p-value). **E.** Median age of cancer accrual according to the classes. The median age is indicated as a dot, and the confidence intervals at 95% are indicated by bars aside the dot. **F.** Proportion of individuals developing more than one cancer during lifetime. For each class, the barplot displays the percentage of individuals with more than one cancer. **G.** Proportion of cancer-free individuals. For each class, the barplot shows the percentage of individuals who did not develop any cancer. **H.** Proportion of individuals with a germline variant in an established cancer predisposing gene (CPG). For each class, the barplot shows the percentage of individuals who carry a variant for a CPG other than *TP53*. **I.** Distribution of clinical classes within *TP53* classes. The proportion of individuals belonging to the following categories are displayed: Li-Fraumeni Syndrome (LFS), Li-Fraumeni like Syndrome (LFL), Chompret criteria (TP53_Chompret), Familial History of cancer (FH), no Familial History of cancer (noFH), Other and Not Applicable (NA). **J.** Distribution of clinical phenotypes of the LFS spectrum definition within *TP53* classes. The proportion of individuals belonging to the categories LFS, attenuated LFS, and incidental LFS are displayed. Not only the P/LP *TP53* variants are included.

Comparison across classes revealed statistically significant differences in tumor patterns (**Figure 4, Table S2, Figures S6 and S7**). Classes 0 and A shared topological and morphological tumor patterns that recapitulated the broad phenotype of the LF spectrum. This included all LFS “signature” cancers such as childhood ACC, CPT, RMS, and MB, teenage OS, and early adulthood breast phyllodes cancers. There were, however, small but statistically significant differences between 0 and A, the latter presenting with a higher proportion of brain cancers (BR) (RR = 1.53, 95% CI [1.16-2.05]), including in particular CPT (**Figure 4B-C, Table S2 and Figure S6**) compared to class 0. This observation suggests that, whereas in most tissues class A variants do not convey a stronger predisposition than class 0 (LOF), they could exert limited dominant GOF effects by accelerating tumor onset in specific tissues (e.g. CPT) without changing the overall phenotypical pattern of LFS. Importantly, we could not identify a subset of variants within class A that would be responsible for these effects (e.g. class A variants overrepresented in CPT compared to other cancers). Overall, these observations suggest that such GOF effects in LFS depend upon cell and tissue context rather than upon specific variant structural and functional properties.

**Figure 4:**
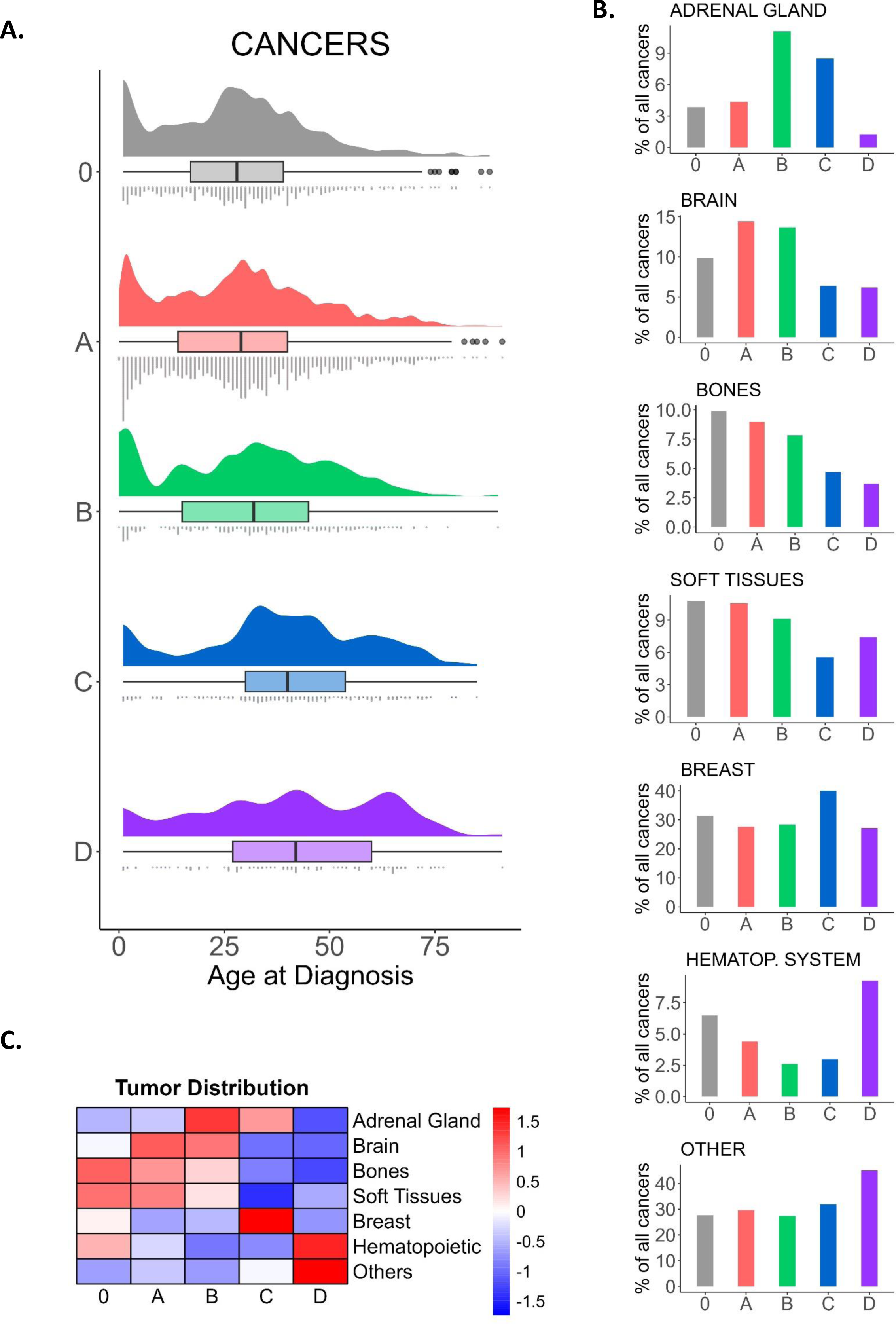
Association of YTA classes with tumor spectrum in the Li-Fraumeni Syndrome. **A.** Age-specific distribution of cancers (all topologies combined) for *TP53* classes. The rain-cloud plots display 1-a density plot showing distribution of age of onset for cancers, 2-a box-plot showing median age of onset as well as quartiles and outliers values, and 3-a dot-plot showing every cancer analyzed. **B.** Distribution of cancers by topology. The most frequent LFS topologies are displayed (adrenal gland, brain, bones, soft tissues, hematopoietic system and breast); all other topologies are referred as “other”. Statistical comparisons are found in Table S2. **C.** Variation of cancer topology within classes. Heatmap synthetizes cancer topology distribution from Figure 4B (normalized in row by cancer topologies).

Compared to 0 and A, class B presented with a slightly smaller proportion of many of the cancers that define the LF spectrum (**Figure 4B-C and Figure S6**), with the notable exception of ACC (RR B vs A = 2.74 (95% CI [1.78;4.143.90]; RR B vs 0 = 3.12 (95% CI [1.84-5.34]) (**Table S2**). In class B, ACC was by far the most common cancer form in children (up to the age of 10), accounting for the fact that cancer accrual in early life was as rapid in that class than in classes 0 or A (**Figure 3C**), despite the relative rarity of some of the other childhood “signature” LFS cancers in class B carriers (i.e., RMS, MB) (**Figure S6**). Compared to A, class B also showed a higher proportion of carriers who were cancer-free (**Figure 3G**) as well as differences in the predominant morphologies of CNS (glioblastomas) (**Figure 4B, Figure S6**). As a result, carriers of class B variants showed a slightly delayed median age of tumor onset as compared to classes 0 and A, but a similar pattern of cancer accrual during childhood. Thus, class B identified variants with a slightly distinct risk profile within the LF spectrum. These variants were mostly distributed within structural motifs of the DBD that are not in contact with DNA and showed less severe structural and functional features than class A. Compared to other classes, class B variants also showed a remarkable heterogeneity in their capacity to transactivate different YTA promoters. Overall, these observations suggest that the more distinct effects of class B variants are due to the retention of partial and selective p53 transcriptional activity. On the other hand, the high prevalence of ACC in carriers of class B variants suggests that perinatal adrenal cortical cells are exquisitely sensitive to wild-type p53 dosage and that variants mediating only partial loss of wild-type p53 activity may suffice to enhance the risk of early ACC.

Class C contains less penetrant variants, which nevertheless predispose to cancers typical of the LF spectrum, in particular ACC (**Figure 4B-C and Figure S6**). Compared to 0, A and B, class C presented with an attenuated risk of all cancers characterizing the LF spectrum, except ACC (RR C vs A = 2.05 (95%CI [1.20-3.34]) and breast cancers (BC) (RR C vs A = 1.75 (95% CI [1.32-2.31]) (**Figure 4B-C, Figure S6, Table S2**). In particular, class C carriers rarely presented with OS compared to class A or 0 carriers (**Figure S6**). Likewise, CPC and RMS were vastly underrepresented, and MB were absent (**Figure S6**). Class C also presented with a higher proportion of cancer-free carriers than any other class (**Figure 3G**). Thus, class C appears to be associated with a cancer risk that only partially recapitulates the LF spectrum. Interestingly, this class included two founder variants qualified as “hypomorphs”, R337H (Brazilian-centric) ^19^ and Y107H (African-centric) ^20^, which retain partial wild-type p53 activities when tested in standard conditions *in vitro* but nevertheless specify a significant risk for diverse cancers of the LF spectrum in both children and adults. It has been suggested that these variants may require synergistic alterations in other pathways to express their full pathogenic potential. This hypothesis is supported by the discovery that, in R337H carriers, an inactivating variant of the putative tumor suppressor *XAF1* (E134*/Glu134Ter/ rs146752602) enhances the risk of developing sarcomas ^21^. Thus, class C may be enriched in incompletely inactivating variants that require genetic, epigenetic, or metabolic/biochemical complementation to express their full pathogenic potential.

Class D presented with the most attenuated pattern within the LF spectrum. “Signature” LF cancers such as early ACC, CPT, MB, OS, and phyllodes tumors of the breast were, if not completely absent, significantly less represented than in A, B or C (**Figure 4B-C, Figure S6**). In contrast, the most represented cancer types in D were malignancies involving the hematopoietic system, the most common form of sporadic neoplasia in children, independent of germline status. Class D variants retained quasi-wild-type p53 transcriptional properties for at least half of the YTA promoters tested, consistent with their ClinVar annotation as benign or likely benign (B/LB). In line with these data, class D carriers also more frequently harbored pathogenic variants in other established cancer predisposition genes than carriers of any other class (**Figure 3H**). These observations suggest that carrying a class D variant may not in itself predispose to LFS. However, it cannot be excluded in specific contexts (e.g. inheritance of other cancer predisposing variants or genetic modifiers), some class D variants may contribute to enhance the risk of LF spectrum cancers. Thus, great care should be exercised when considering such variants in genetic counselling.

The overall cancer phenotypes of each variant class are summarized in a heatmap (**Figure 4C**), highlighting an LF tumor-specific phenotypic gradient from classes 0 and A to class C. Classes A and 0 were associated with the most severe forms of LFS, class B identified a distinct, slightly less penetrant form, with ACC as the most frequent early life cancer, class C was correlated to an attenuated form nevertheless predisposing to cancers typical of the LF spectrum (ACC, BC), whereas class D did not appear to confer a risk for LFS cancers.

### Dissecting variant classes using cell-based functional scores

Despite their high sensitivity and selectivity for distinct p53RE, YTA screens may lack the specificity of phenotypic screens in human cells addressing physiologically meaningful p53 functionalities. We thus sought to determine whether the functional scores obtained in systematic human cell-based assays were consistent with our YTA classes and could identify additional groups of variants associated with distinct LF phenotypes. First, the stability and coherence of YTA classes was challenged with the functional scores developed by Giacomelli et al. ^11^ (phenotypic selection model) and Kotler et al. ^13^ (relative fitness score). **Figure 5** shows that these functional scores were broadly consistent with YTA classes, with A having the highest and D the lowest median scores. However, individual variant scores were widely distributed within each class, suggesting that YTA classes may contain variants that differ by their functional properties when overexpressed in human cells. To further assess this heterogeneity, we separated variants in each class into groups according to the quartiles of their human cell-based functional scores (**Figures S8, S9**). Results show that cancer accrual in class A carriers was similar across all quartiles and thus independent of human cell-based scores. Whereas heterogeneity was found for classes B, C and D, it reached statistical significance only for class C (p < 0.0001, Giacomelli’s phenotypic selection model; p = 0.0073, Kotler’s relative fitness score), highlighting that this class contains variants that are more functionally heterogeneous than variants in other classes. Similarly, stability analyses using TP53_PROF, an integrative computational score ^17^, did not show any additional benefit of using this score for stratifying variants according to their pathogenicity within each of the YTA classes (**Figure S10**). Overall, these results show that YTA classes were remarkably robust and sensitive in predicting cancer accrual and cancer phenotypes in germline *TP53* variant carriers.

**Figure 5:**
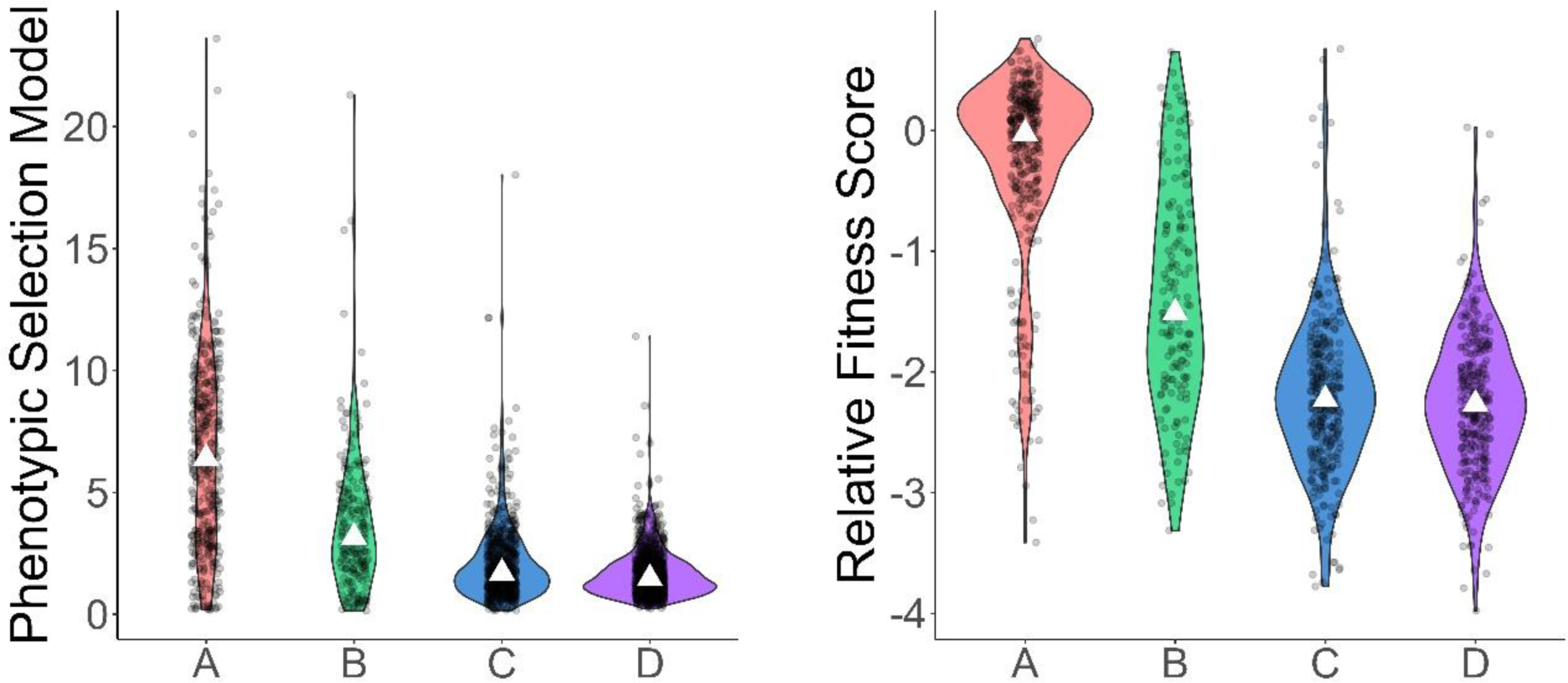
Heterogeneity of *TP53* scores within YTA classes. Distribution of functional scorings (phenotypic selection model and relative fitness score) into all 2,314 *TP53* missense variants, subdivided by YTA classes. Violin plots and dot-plots display the distribution of scores, and the median scores are indicated as white triangles.

### Clinical significance of YTA classes

ClinVar expert panel annotations (https://clinicalgenome.org/affiliation/50013/) are based on expert consensus review of genotype/phenotype correlations and are the current standard for interpreting *TP53* germline variants for clinical purposes ^14^. However, these annotations cover only a proportion of all missense variants in the NCI germline dataset. We thus assessed the concordance between YTA classes and ClinVar expert panel as a first step to determine whether YTA classes could assist in the clinical evaluation of variants that have not yet been annotated to date. **Figure 6A** shows that YTA classes were broadly concordant with expert panel annotations, with A and B classes mostly represented among P/PL variants and C and D classes mostly represented among B/LB variants. The only exception was D49H, a class A variant which was annotated as LB by ClinVar. Of note, there is only a single family (2 patients) with this variant documented in the NCI germline dataset. This family matched phenotypic criteria for TP53 mutation testing (Chompret’s criteria; embryonal RMS of the cervix at 2 years and Hodgkin’s lymphoma at 15 years) ^22^. D49H has also been reported in 6 Japanese patients with family history of cancer, one of whom matched Chompret’s criteria ^23^. This patient carried another germline class A TP53 variant, A159D. Overall, these observations suggest that D49H should be considered with caution as a rare, potentially pathogenic variant.

**Figure 6:**
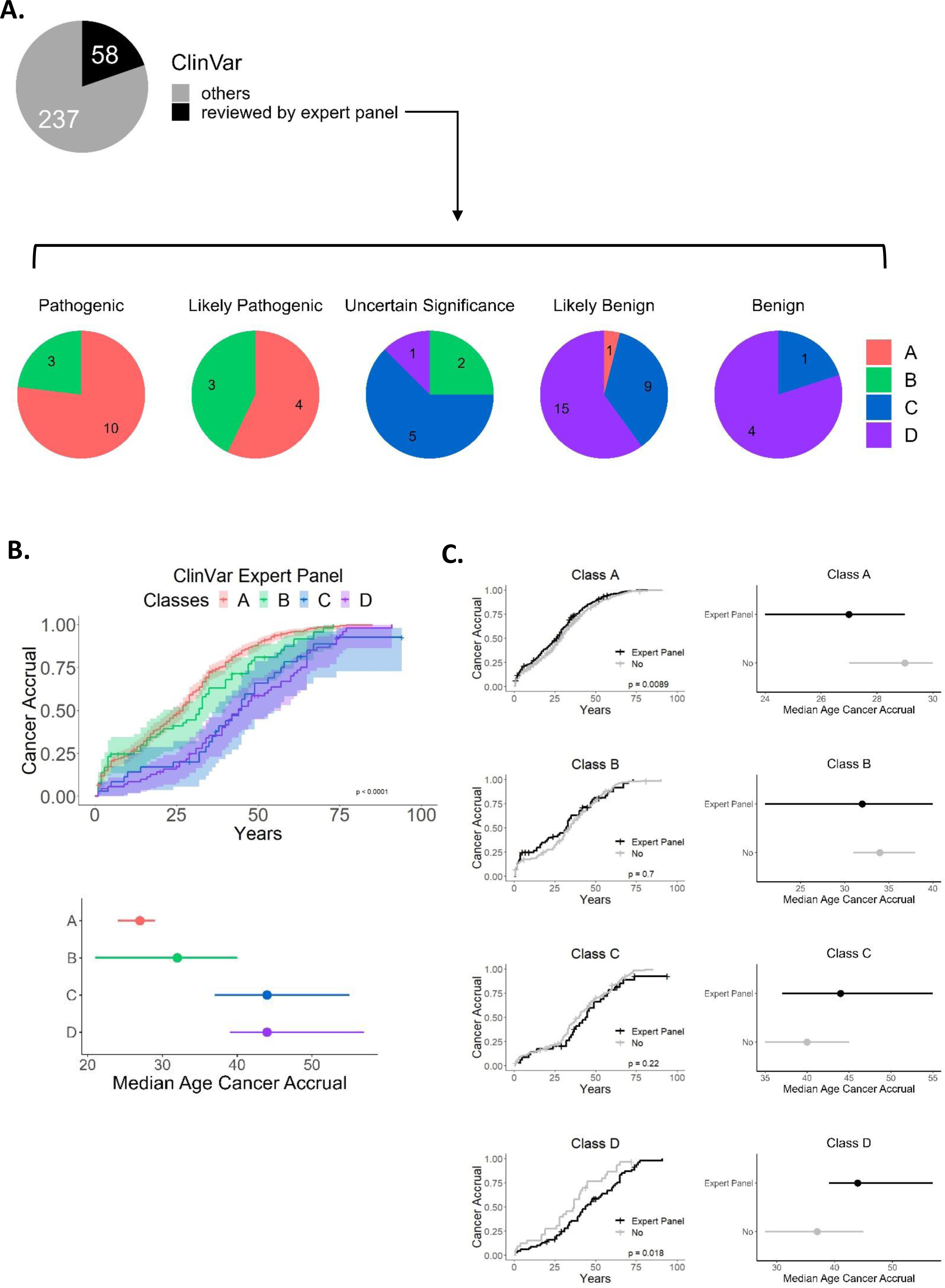
Concordance between YTA classes and ClinVar annotations. **A.** Distribution of expert panel reviewed and non-expert panel reviewed *TP53* variants in the ClinVar classification. Variants from the IARC/NCI *TP53* germline database are indicated in the pie chart. The second row of pie charts represents the breakdown of expert panel reviewed variants in ClinVar categories (Pathogenic, Likely Pathogenic, Uncertain Significance, Likely Benign and Benign). Mapping of the YTA classes within each ClinVar category is displayed. The number of variants for each ClinVar category is indicated within each pie chart. **B.** Cancer accrual of each YTA class for the subcategory of expert panel reviewed *TP53* variants (top panel). The first cancer of the NCI/IARC TP53 germline database is used to monitor cancer accrual. Confidence intervals at 95% and Log-Rank test p-value are indicated. Median age of cancer accrual according to the classes (bottom panel). The median age is indicated as a dot, and the confidence intervals at 95% are indicated by bars aside the dot. **C.** Comparison of cancer accrual for variants annotated by expert panel versus variants not annotated by expert panel (No). Left panels show cancer accrual within each YTA class. Right panels display the median and confidence intervals.

Interestingly, variants annotated as “Uncertain Significance” (VUS) were predominantly class C variants, followed by class B and D variants, suggesting that YTA classes may help in resolving ambiguities in variant annotations. Furthermore, variants annotated by the expert panel could be perfectly separated on the basis of cancer accrual according to YTA classes (p < 0.0001), recapitulating the distinct accrual pattern of each class (e.g the difference in cancer accrual between class A and class B variant carriers). (**Figure 6B and 3C**). These results suggest that YTA classes are remarkably consistent with expert panel annotations.

Next, we evaluated to which extend YTA classes could predict cancer accrual in subjects carrying variants not annotated by the expert panel. Within classes A, B and C, cancer accrual in carriers of non-expert panel variants was almost perfectly aligned with the one of expert panel curated variants (**Figure 6C**).

To further assess the usefulness of YTA classes in a clinical context, we examined cancer accrual and tumor patterns according to classes in three cohorts of carriers recruited in different high risk cancer predisposition clinics in Germany (n = 146, LFS Registry in Hannover), France (n = 578, French LFS Cohort) and Canada (n = 97, Toronto LFS Cohort). These cohorts are maintained and annotated mainly independently of the NCI germline dataset. **Figure 7** shows that, in each of the cohorts, YTA classes correctly predicted cancer accrual and distinguished among carriers with quantitative and qualitative differences in LF spectrum. In all three cohorts, class A carriers presented the most severe phenotype (similar to class 0 in the German and Canadian cohort). Compared to A, class B carriers had a distinct and less penetrant phenotype, characterized by high risk of early life ACC and slightly delayed overall cancer accrual. Class C carriers often had attenuated phenotypes, nevertheless including cancers typical of the LF spectrum (i.e., ACC and PBC) but excluding most other typical LF childhood cancers, whereas in class D tumor patterns tended to be more heterogeneous, with the majority of carriers not matching LF spectrum definitions (**Figure S11**). Overall, these observations support that YTA classes can help to predict specific patterns of risk in germline carriers and could be particularly helpful for assessing the risk of novel variants as well as variants not yet annotated by expert panels or ClinVar.

**Figure 7:**
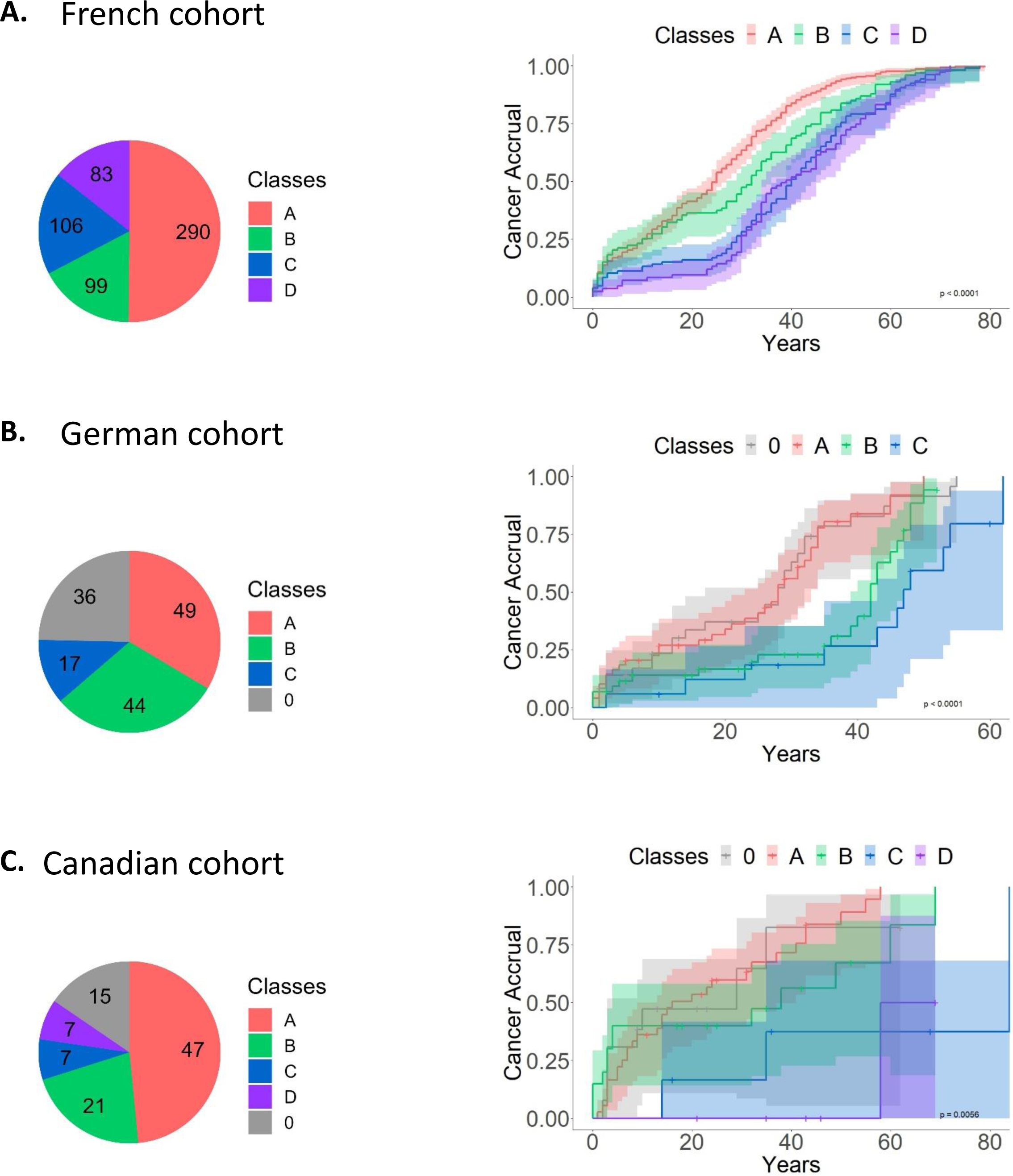
Matching YTA classes to Li-Fraumeni clinical validation cohorts. For each cohort, the patients’ distribution into *TP53* classes are shown in the pie charts (left panels), as well as the cancer accrual using the age of onset for the first cancer, confidence intervals at 95% and Log-Rank p-values (right panels). **A.** Cohort 1 (France) analysis. **B.** Cohort 2 (Germany) analysis. **C.** Cohort 3 (Canada) analysis.

## CONCLUSIONS

This new classification of *TP53* missense variants into 4 classes supports a robust interpolation between genotypes (variant characteristics) and phenotypes (cancer risk and spectrum). The classification is based on quantitative and qualitative similarities between variants in a yeast-based transcriptional assay. Whereas the yeast readout is based on variants’ interaction with synthetic p53RE (DNA binding capacity), YTA classes correctly separate severe from mild or benign variants in all structural domains of the p53 protein, including transactivation domain, DBD and tetramerization domain. The coherence and robustness of YTA classes is demonstrated by the fact that implementing human cell-based functional scores within YTA classes did not break them into further subgroups, suggesting that yeast-based readouts are precise and sensitive enough by themselves to allocate variants to clinically meaningful groups. Overall, these observations imply that, collectively, the 8 yeast-based readouts available for each variant can be used as sensitive “mugshots” for variant risk assessment.

This classification provides new insights into the concept of attenuation within the LF spectrum ^5^. The term aLFS has been coined to identify an LF phenotype defined by the presence of a germline P/LP *TP53* variant in a person with any cancer who does not meet LFS genetic testing criteria and has no cancer diagnosed before age 18 years. Our analysis did not identify a specific YTA class matching this phenotypic definition. Rather, we observed a gradient in the proportion of individuals with aLFS phenotype from class 0 or A (< 17-21%) to class C or D (> 50-61%) (**Figure 3J**). Remarkably, even among carriers who met LFS genetic testing criteria, YTA classes underscored a gradient of phenotypic attenuation, with carriers of B or C variants presenting, on average, with a less severe phenotype than carriers of A or 0 variants. Another key message is that risk attenuation in relation to YTA classes does not obey to a unique, linear rule for all cancers that characterize the LF spectrum. In particular, carriers of B or C variants retain a risk for early ACC at least equal to classes 0 and A, supporting that predisposition to this cancer can be high even with relatively mild *TP53* variants. In contrast, other “signature” LF cancers such as OS, CPT, MB or RMS, were remarkably more prevalent in carriers of 0 or A than in carriers of B, C or D variant classes. A limitation of this study is that it does not take into account the fact that at least some mutations causing missense variants may also cause aberrant mRNA splicing patterns, resulting in loss of p53 protein expression. Such effects may confer greater severity to variants currently solely interpreted as missense. So far, these effects on splicing have not been evaluated in systematic functional assays. Overall, our integrative genotypic/phenotypic analysis provides a new angle to predict individual and familial risk in germline variant carriers, as well as a basis to identify hypomorphic variants that require genetic or epigenetic modification to express their full pathogenic potential.

## MATERIAL AND METHODS

### Iterative clustering on transactivation data

We retrieved transactivation scores from 2,314 *TP53* variants ^12^ on eight *TP53* promoters (WAF1nWT, MDM2nWT, BAXnWT, h1433snWT, AIP1nWT, GADD45nWT, NOXAnWT, P53R2nWT). We performed hierarchical Ward clustering in R v3.6.1 with Euclidean distance, and arbitrarily chose three clusters. We performed a second iteration of clustering in the sub-clusters including more than 50 mutations. Up to three layers of clustering iterations were processed. A total of 16 distinct clusters were thus defined. Dendrograms were retrieved with *dendextend* package, and heatmaps were built using *pheatmap* package.

### YTA classes consolidation

Final groups of clusters (YTA classes) were defined via the following approach: the most disrupted variants (cluster 1_1_1) were assigned to class A; variants of the upper layer (layer 1_1 except cluster 1_1_1) were defined as class B; variants of the next upper layer (layer 1 except the layer 1_1) were defined as class C; finally, the less disrupted variants (all variants except layer 1) were defined as class D.

### Confrontation of YTA classes with existing scores for *TP53*

We confronted our classes to various scores attributed to *TP53*: the transactivation classes as previously defined by the analysis of transactivation in yeast assays ^12^, the Sorting Intolerant from Tolerant (SIFT) database (https://sift.bii.a-star.edu.sg) ^16^, the Align Grantham Variation Grantham Deviation (AGVGD) database (http://agvgd.hci.utah.edu) ^15^, the TP53 prediction of functionality (TP53_PROF) ^17^, the *TP53* functionality scores obtained in human cell lines ^11,13^ and the ClinVar database which aggregates information about genomic variation and its relationship to human health (https://www.ncbi.nlm.nih.gov/clinvar) ^24^. The confrontation of the classes with the various *TP53* scores was performed using Pareto Plot or Pie Chart representations for categorical values and Scattered Plots and Violin Plots for numerical values (R, *ggplot2* package).

### Structural mapping of YTA classes

The distribution of *TP53* mutated residues along the TP53 sequence and domains was analyzed for each YTA class. Heatmaps were used to highlight the pattern of distribution of variants along *TP53* sequence in the different YTA classes (R, package *pheatmap*). The proportion of variants within each *TP53* domain is depicted using cumulative histograms (R, package *ggplot2*). The localization of mutations in the p53 protein structure was assessed using the software ChimeraX ^25^ (https://www.cgl.ucsf.edu/chimerax) together with PDB files of the *TP53* binding domain 3KMD ^26^ and the *TP53* tetramerization domain 1C26 ^27^, as well as the alpha-fold modelized structure ^28^.

### YTA classes distribution in datasets

The occurrence of *TP53* variants assigned to YTA classes was assessed in different databases: tumor-based variants in the Cosmic database v97 (https://cancer.sanger.ac.uk/cosmic) ^29^, normal sample-based variants in the gnomAD database v3.1.2 (https://gnomad.broadinstitute.org) ^30^, and *TP53* germline variants database R20 released in July 2019 and hosted at the NCI (https://tp53.isb-cgc.org) ^18^. We used the protein mutation information in the databases to analyze the distribution of variants into YTA classes. Pie charts are used to show distribution of variant classes in the different databases, and numerical counts are indicated (R, *ggplot2* package).

### Selection of individuals from the LF spectrum database

We analyzed the YTA classes on clinical data from the LF spectrum database R20 (germline *TP53* variants), released in July 2019 and hosted at the NCI (https://tp53.isb-cgc.org) ^18^. This database recapitulates clinical data of patients carrying *TP53* germline variants. The database contains data from 3,446 individuals (from 1,522 families) reported in the scientific literature since 1990. Individuals carrying several *TP53* variants were excluded from our analysis because of complexity to determine the contribution of individual variants in these patients. Also, individuals carrying the *TP53* p.R337H Brazilian variant who were not recruited through familial history of cancer were removed, because of bias of analysis (in column Ref_ID: 138, 196, 259, 323 and 400 were selected in the LF spectrum database). Individuals carrying *TP53* missense variants were dispatched in groups corresponding to the YTA classes (A, B, C, D). The remaining individuals, carrying non-missense variants were selected based on the type of effect of the variant in order to build a class of reference containing non-functional (LOF) variants. We only kept nonsense and frameshift (ft) variants (from column “Effect”) to consolidate a null class (named class 0) corresponding to disruption of the full-length protein.

### Genotype-Phenotype correlations of YTA classes within the LF spectrum database

Lifetime cancer accrual for the YTA classes was assessed using the age of first cancer onset or the cancer-free age of an individual. Patients without age indication were not considered for this analysis. The age of median cancer accrual (and confidence intervals at 95%) was extracted and a Log-Rank Mantel-Cox test was run to assess the significance of differences between groups. These analyses were performed using R and the *survival* and *survminer* packages. Information about individuals’ sex was retrieved in the corresponding column (Sex). The proportion of individuals with multiple cancers versus individuals with one cancer was assessed for each YTA class. The proportion of cancer-free individuals compared to individuals with cancer was assessed for each class. Tumor distribution was determined in the YTA classes by analyzing all tumors described in the dataset (primary and secondary malignancies). We used the topology (organs) and the morphology (subtypes) information available in the LF spectrum database, and we consolidated groups based on organs of interest for the study of LFS (adrenal gland, brain, bone, soft tissue, breast, hematopoietic system, and the mix of all other organs) as well as specific cancer types (ACC, OS, CPT, MB, glioblastoma, phyllodes tumors, and RMS). Graphical representation of proportions of tumor types was performed using R (package *ggplot2*). Distribution of tumor topology was compared between YTA classes by performing a Khi2 test (multiple pairwise comparison) to obtain a risk ratio, confidence intervals at 95% and an associated adjusted p-value, using R software. Mapping of age-specific tumor distribution within YTA classes was assessed by generating rain-cloud plots (R, package *ggplot2*). Finally, the relationship between YTA classes and the clinical definitions of LFS was evaluated using the LFS spectrum classification ^5^ and the column “Class” from the IARC/NCI database, to build bar-plots of proportions of individuals within each clinical class.

### Challenging YTA classes with other functional scores

YTA classes were separated based on the *TP53* functionality scores obtained in human cell lines ^11,13^. For each YTA class, *TP53* variants were separated into four groups by quartile values. The 1^st^ quartile corresponds to lower scores and the 4^th^ quartile corresponds to higher scores (most disrupted functionality of p53). YTA classes were also separated based on the *TP53* prediction of functionality (TP53_PROF, Deleterious and Non-Deleterious) ^17^ and the ClinVar database annotation (https://www.ncbi.nlm.nih.gov/clinvar) ^24^. Cancer accrual of the subgroups of each YTA class was analyzed and the median age and statistical differences were extracted similar to YTA class comparison.

### Validation cohorts

Three validation cohorts originating from three different clinics were used to verify the consistency of our observations for the YTA classes.

- The “LFS Registry in Hannover”, Germany, recruits patients with a previously established diagnosis of LFS. Only patients carrying pathogenic or likely pathogenic variants (according to Fortuno criteria) are included.
- The “French LFS Cohort” has included germline missense TP53 variants from the French registry (Rouen, France) identified in cancer patients who benefited from TP53 analysis.
- The “Toronto LFS Cohort”, Canada, is a multi-institutional collection of data from patients and families carrying germline pathogenic variants that were referred to The Hospital for Sick Children.

## Data Availability

All data produced in the present study are available upon reasonable request to the authors

## AUTHOR CONTRIBUTIONS

EM, NL and PH conceived and designed the study. PH supervised research. NL performed hierarchical clustering. EM performed bioinformatic analysis and graphical representations with the help of SB. EM analyzed genotype-phenotype correlations with the help of CF. JP, CK, GB, NF and DM provided clinical data. AA and FC assisted EM with statistical analyses. JP, CK, GB, NF, DM, MIA, AL and CG contributed to the discussion and interpretation of variant classes. EM and PH wrote the manuscript. All authors critically revised the manuscript for important intellectual content and approved the version for publication.

## ACKNOLEDGMENTS

EM was a recipient of a European MSCA individual fellowship (846806) and a Foundation ARC (France) postdoctoral fellowship. This work is supported by the IDEX of University Grenoble Alpes (LIFE project) and by the ERiCAN program of Foundation MSDAvenir (France). CPK is supported by the BMBF ADDRess (01GM2205A) and by the Deutsche Kinderkrebsstiftung (DKS2021.25).

## ABBREVIATIONS

ACC: Adrenal Cortical Carcinoma
AGVGD: Align Grantham Variation and Grantham Deviation
aLFS: attenuated LFS
BC: Breast Cancer
BR: Brain Cancer
B/LB: Benign or Likely Benign
CNS: Central Nervous System Tumors
CRC: Colorectal Cancer
COSMIC: Catalogue of Somatic Mutations in Cancer
CPT: Choroid Plexus Tumors
DBD: DNA Binding Domain
DNE: dominant-negative effect
GOF: Gain of Function
gnomAD: Genome Aggregation Database
LF: Li-Fraumeni
LFS: Li-Fraumeni Syndrome
LOF: loss of function
LUAD: lung adenocarcinomas
MB: Medulloblastoma
OS: Osteosarcoma
PBC: Premenopausal Breast Cancer
P/LP: Pathogenic or Likely Pathogenic
P53RE: p53 Response Elements
RMS: Rhabdomyosarcoma
SIFT: Sorting Intolerant From Tolerant
STS: Soft Tissue Sarcoma
VUS: Variants of Unknown Significance
YTA: Yeast Transactivation Assay

## SUPPLEMENTARY FIGURE AND TABLE LEGENDS

**Figure S1: Yeast Transactivation Assay (YTA) -based iterative clustering of the *TP53* missense variants.** Transcriptional data from yeast-based assays consisting of eight measurements for 2,314 *TP53* missense variants were retrieved from Kato et al. Three layers of Ward hierarchical clustering were applied to separate variants. For each layer, heatmaps display the transactivation scores, and dendrograms represent the clusters defined by hierarchical clustering.

**Figure S2: Hierarchy of clusters and consolidation of *TP53* variants classes. A.** Distribution of the 2,314 *TP53* missense variants in clusters. Numbers of variants are indicated in parenthesis above the cluster name. Color codes correspond to the consolidation of classes explained in Fig S2B. **B.** Consolidation of classes. Class A variants correspond to cluster 1_1_1; Class B variants correspond to 1_1 cluster except class A; Class C variants correspond to cluster 1 except classes A and B; Class D variants correspond to all variants except classes A, B, and C. **C.** Distribution of transactivation scores within the consolidated classes. The heatmap displays scores for all eight promoter measurements regarding all *TP53* variants ranked by YTA classes. **D.** Distribution of transactivation classes (non-functional, partially functional, functional and supertrans) of *TP53* variants within YTA classes based on hierarchical clustering. Pareto plots display the number of *TP53* variants belonging to the transactivation classes.

**Figure S3: Structural localization of variants from YTA classes. A.** *TP53* missense variants located on Alpha-Fold *TP53* structural modeling. The TP53 germline variants found in the IARC/NCI LFS database are separated by YTA classes and displayed on the 3D structure. **B.** *TP53* missense variants located on crystal-based structures of the DNA binding domain (top panel) and oligomerization domain (bottom panel). *TP53* germline variants found in the IARC/NCI LFS database are separated by YTA classes and displayed on the 3D structure.

**Figure S4: Mapping of YTA classes to *TP53* scores.** Pareto plots showing the distribution of functional scorings for *TP53* variants (SIFT, AGVGD, and TP53 PROF) within YTA classes.

**Figure S5: Designing the *TP53* variants class 0. A.** Cancer accrual for all categories of non-missense variants in individuals from the IARC/NCI *TP53* variants germline database (LFS). **B.** Comparison of median age of cancer accrual (red dots) and confidence intervals (black lines) for the categories of non-missense variants. The nonsense and frameshift variants are selected for the design of a class 0.

**Figure S6: Association of YTA classes with tumor spectrum in LFS.** Distribution of cancers by morphology subtype. Histograms displaying the percentage of all cancers of individuals from IARC/NCI *TP53* variants germline database (LFS) with specific tumor morphologies evoking LFS.

**Figure S7: YTA class specificities in the distribution of different topologies of cancers by age.** Rain-cloud plots showing the age distribution of the following cancer topologies: adrenal gland, bones, breast, soft tissues, brain, hematopoietic, and all other topologies (other), for the individuals of classes 0, A, B and C in the IARC/NCI *TP53* variants germline database (LFS). Class D was not considered as counts of cancers were not sufficient for analysis.

**Figure S8: Challenging YTA classes with the phenotypic selection model score in LFS. A.** Distribution of the phenotypic selection model score from Giacomelli et al. for each variant from the IARC/NCI *TP53* variants germline database. For each YTA class, dot-plots are annotated with the corresponding quartile values (25%; median; 75%) to design four groups (quartile 1, quartile 2, quartile 3, and quartile 4, from lowest to highest values). **B.** Comparison of cancer accrual for individuals belonging to the four quartile groups in the same YTA class (left panels), and relationship between median age of cancer accrual and quartiles (right panel).

**Figure S9: Challenging YTA classes with the relative fitness score in LFS. A.** Distribution of the relative fitness score from Kotler et al. for each variant from the IARC/NCI *TP53* variants germline database. For each YTA class, dot-plots are annotated with the corresponding quartile values (25%; median; 75%) to design four groups (quartile 1, quartile 2, quartile 3, and quartile 4, from lowest to highest values). **B.** Comparison of cancer accrual for individuals belonging to the four quartile groups in the same YTA class (left panels), and relationship between median age of cancer accrual and quartiles (right panel).

**Figure S10: Mapping YTA classes to the *TP53* PROF classification. A.** Distribution of YTA classes in deleterious (D) and non-deleterious (ND) classes of the TP53_PROF model from Ben-Cohen et al. The number of variants from the *TP53* germline database NCI/IARC is displayed. **B.** Cancer accrual of individuals separated by YTA classes, for deleterious and for non-deleterious *TP53* variants. Log-Rank p-values are indicated.

**Figure S11: Association of YTA classes with tumor spectrum in Li-Fraumeni clinical validation cohorts.** Distribution of cancers by topology. The most frequent LFS topologies are displayed (adrenal gland, brain, bones, soft tissues, hematopoietic system and breast); all other topologies are referred as “other”; cancer-free patients are indicated as “no cancer”. **A.** Cohort 1 (France) analysis. **B.** Cohort 2 (Germany) analysis. **C.** Cohort 3 (Canada) analysis.

**Table S1: *TP53* missense variants and corresponding YTA classes.** The 2,314 *TP53* missense variants are annotated with the YTA classification (classes A, B, C, D), as well as scores developed by others.

**Table S2: Risk ratio of tumor localization for YTA classes in LFS.** Statistical significance of variation of tumor distribution (topology) for each pair-wise comparison of YTA classes. Khi2 tests are reported in the table (risk ratio between the two classes, with confidence intervals 95% and the adjusted p-value with Benjamini-Hochberg correction for multiple comparisons), for the following topologies: adrenal gland, bones, brain, breast, hematopoietic system, other, and soft tissues. Grey highlights correspond to pairwise comparison that reached adjusted p-value <0.05.

## Notes

### Competing Interest Statement

The authors have declared no competing interest.

### Funding Statement

This study was funded by:
- European Commission (MSCA IF 846806)
- Fondation ARC
- University Grenoble Alpes (IDEX LIFE)
- Fondation MSD Avenir (ERICAN)
- BMBF ADDRess (01GM2205A)
- Deutsche Kinderkrebsstiftung (DKS2021.25)

### Author Declarations

https://tp53.isb-cgc.org/ https://cancer.sanger.ac.uk/cosmic https://gnomad.broadinstitute.org https://www.ncbi.nlm.nih.gov/clinvar https://doi.org/10.1136/jmedgenet-2017-104976 https://doi.org/10.1186/s13045-022-01332-1 https://doi.org/10.1158/2767-9764.CRC-22-0402

