## Supplementary Figures for "Fine resolution clustering of *TP53* variants into functional classes predicts cancer risks and spectra among germline variant carriers"

**Affiliations:**
(1) Univ. Grenoble Alpes, Inserm 1209, CNRS 5309, Institute for Advanced Biosciences, F38000, Grenoble, France
(2) Pediatric Hematology and Oncology, Hannover Medical School, Hannover, Germany
(3) Centre Hospitalier Universitaire Grenoble Alpes, Department of Pediatrics
(4) Department of Pediatrics, Division of Hematology-Oncology, Montreal Children's Hospital, McGill University Health Centre, Montreal, Quebec, Canada
(5) Department of Prevention Cancer Environment, Centre Léon Bérard, Lyon, France
(6) Genetics and Genome Biology Program, The Hospital for Sick Children; University of Toronto, Toronto, Ontario, Canada
(7) Department of Oncology, Hospital Sírio-Libanês, São Paulo, Brazil
(8) Simons Center for Systems Biology, Institute for Advanced Study, Princeton, NJ
(9) Department of Genetics, Normandy Center for Genomic and Personalized Medicine, University Hospital, Rouen, France; Normandie Univ - UniRouen, Inserm U1245, Rouen, France

**Correspondence:**
* corresponding author:
Institute for Advanced Biosciences,
Univ. Grenoble Alpes, Inserm 1209, CNRS 5309
Site Santé, Allée des Alpes
F38700, La Tronche, France


#### ABSTRACT

Li-Fraumeni syndrome (LFS) is a heterogeneous predisposition to a broad spectrum of cancers caused by pathogenic *TP53* germline variants. We have used a clustering approach to assign missense variants to functional classes with distinct quantitative and qualitative features based on transcriptional activity in yeast assays. Genotype-phenotype correlations were analyzed using the germline *TP53* mutation database (n = 3,446) and validated in three LFS clinical cohorts (n= 821). Carriers of class A variants recapitulated all traits of fully penetrant LFS (median age at first diagnosis = 28 years). Class B carriers showed a less penetrant form (median = 33 years, $p < 0.05$) dominated by adrenocortical and breast cancers. Class C or D carriers had attenuated phenotypes (median = 41 years, $p < 0.001$) with typical LFS cancers in C and mostly non-LFS cancers in D. This new classification provides insight into structural/functional features causing pathogenicity.

**Figure S1**

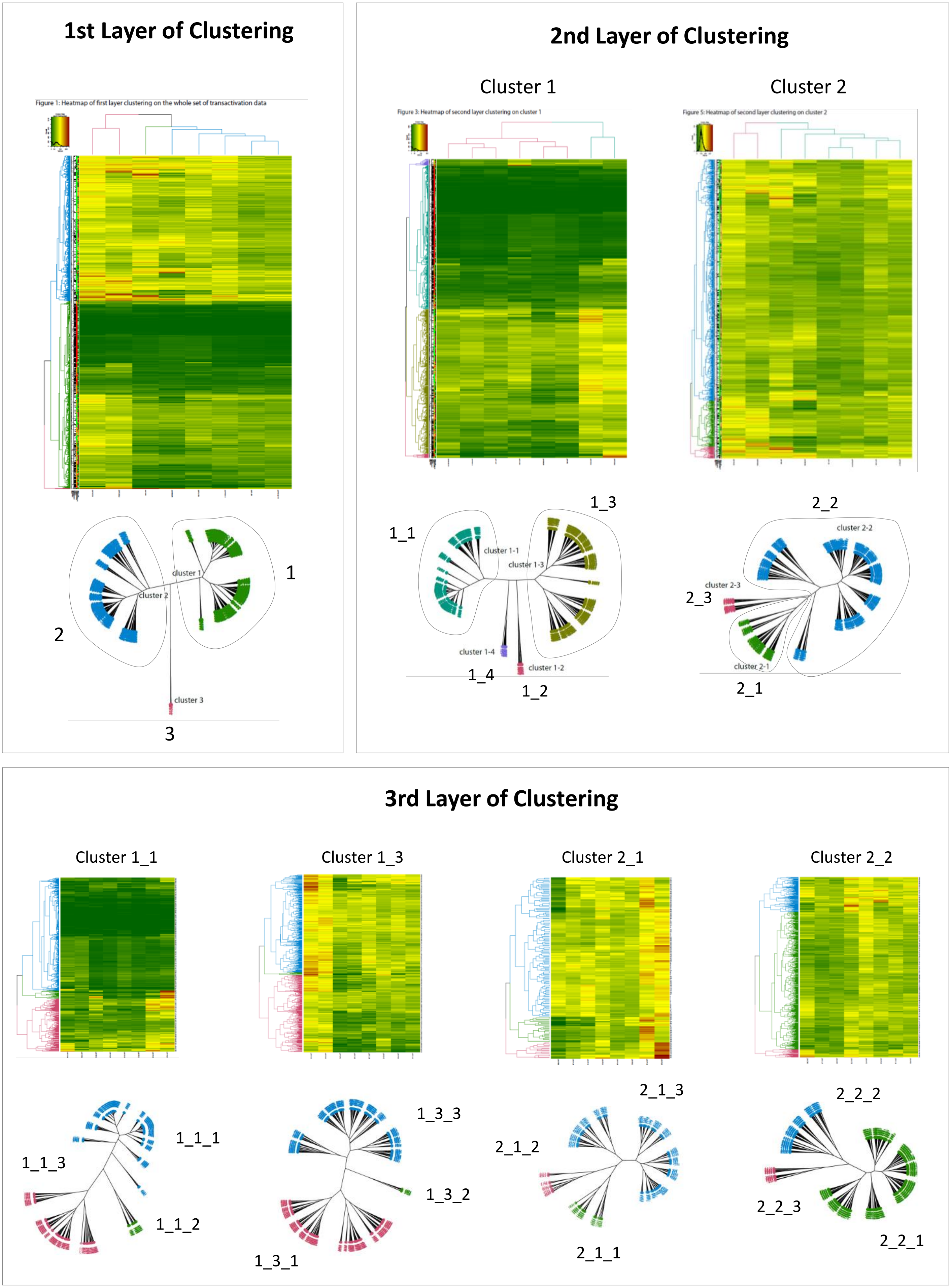
1st Layer of Clustering
Figure 1: Heatmap of first layer clustering on the whole set of transactivation data
cluster 1
cluster 2
cluster 3
1
2
3
2nd Layer of Clustering
Cluster 1
Cluster 2
Figure 3: Heatmap of second layer clustering on cluster 1
Figure 5: Heatmap of second layer clustering on cluster 2
1_1
1_3
cluster 1-1
cluster 1-3
cluster 1-4
cluster 1-2
1_4
1_2
2_2
cluster 2-2
cluster 2-3
2_3
cluster 2-1
2_1
3rd Layer of Clustering
Cluster 1_1
Cluster 1_3
Cluster 2_1
Cluster 2_2
1_1_1
1_1_3
1_1_2
1_3_3
1_3_2
1_3_1
2_1_3
2_1_2
2_1_1
2_2_2
2_2_3
2_2_1

#### Figure S2

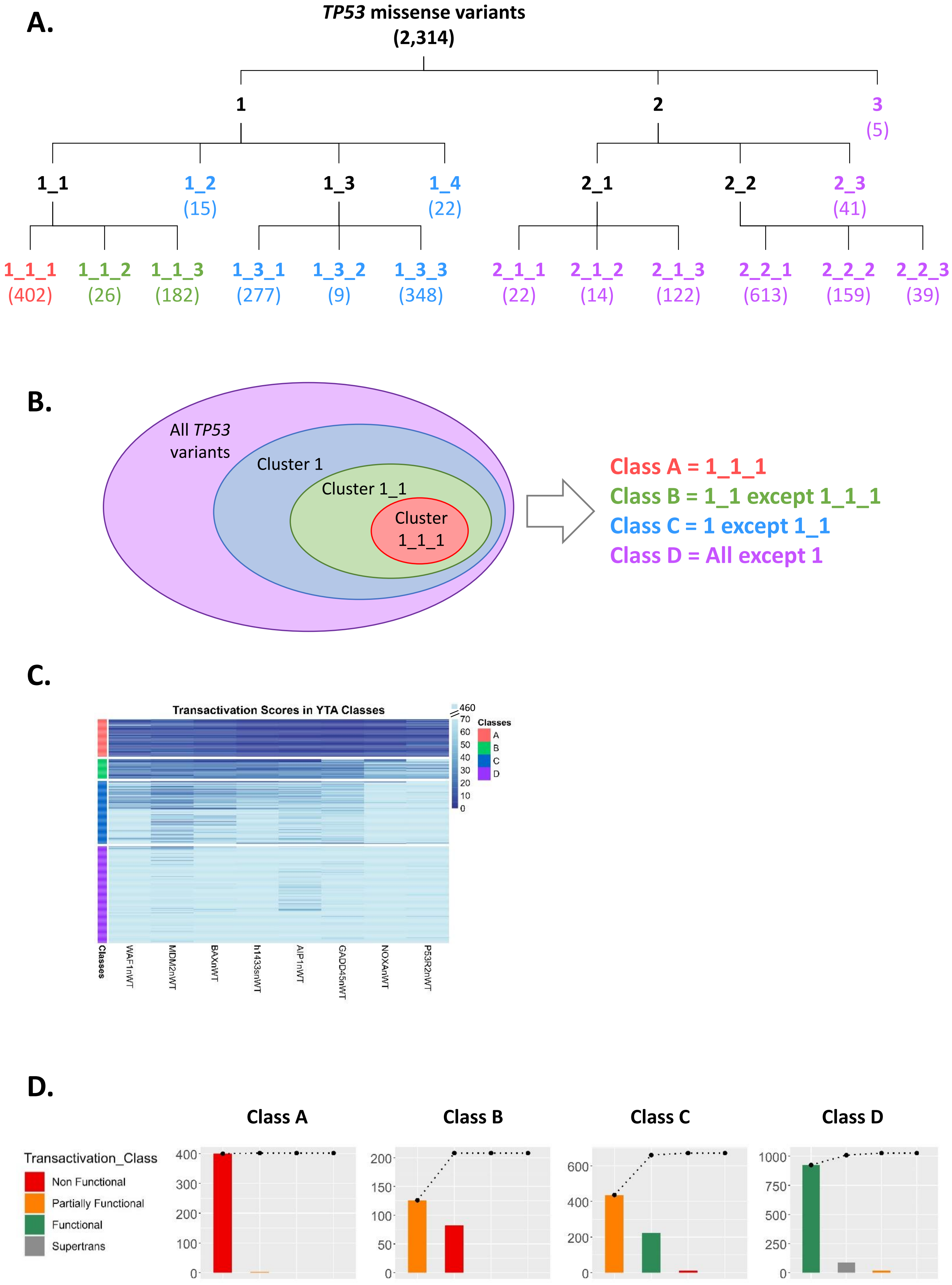
A.
TP53 missense variants
(2,314)
1
2
3
(5)
1_1
1_2
(15)
1_3
1_4
(22)
2_1
2_2
2_3
(41)
1_1_1
(402)
1_1_2
(26)
1_1_3
(182)
1_3_1
(277)
1_3_2
(9)
1_3_3
(348)
2_1_1
(22)
2_1_2
(14)
2_1_3
(122)
2_2_1
(613)
2_2_2
(159)
2_2_3
(39)
B.
All TP53 variants
Cluster 1
Cluster 1_1
Cluster 1_1_1
Class A = 1_1_1
Class B = 1_1 except 1_1_1
Class C = 1 except 1_1
Class D = All except 1
C.
Transactivation Scores in YTA Classes
460
70
60
50
40
30
20
10
0
Classes
A
B
C
D
Classes
WAF1nWT
MDM2nWT
BAXnWT
h1433snWT
AIP1nWT
GADD45nWT
NOXAnWT
P53R2nWT
D.
Class A
Class B
Class C
Class D
Transactivation_Class
Non Functional
Partially Functional
Functional
Supertrans
400
300
200
100
0
200
150
100
50
0
600
400
200
0
1000
750
500
250
0

### Figure S3

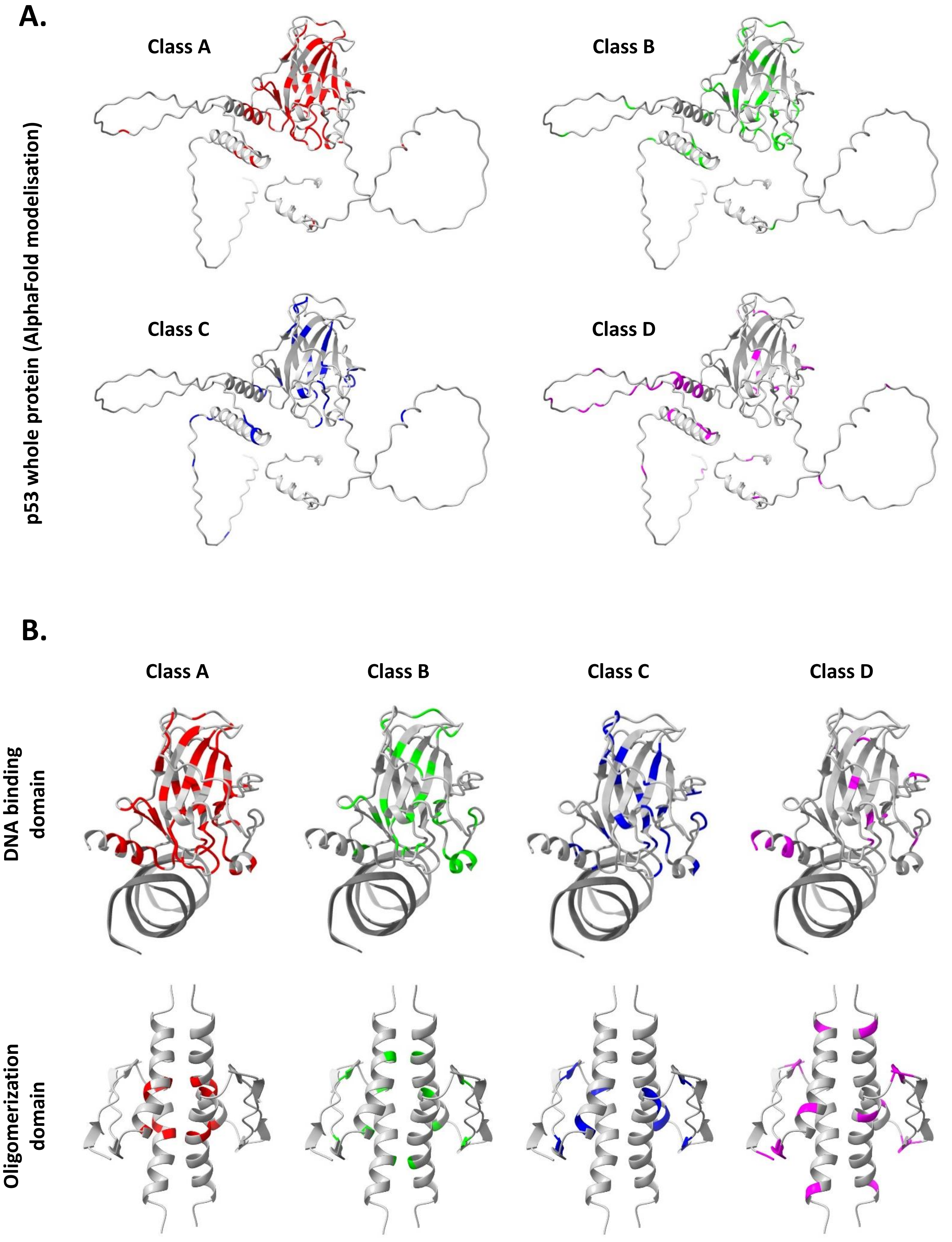
A.
Class A
Class B
Class C
Class D
p53 whole protein (AlphaFold modelisation)
B.
Class A
Class B
Class C
Class D
DNA binding domain
Oligomerization domain

### Figure S4

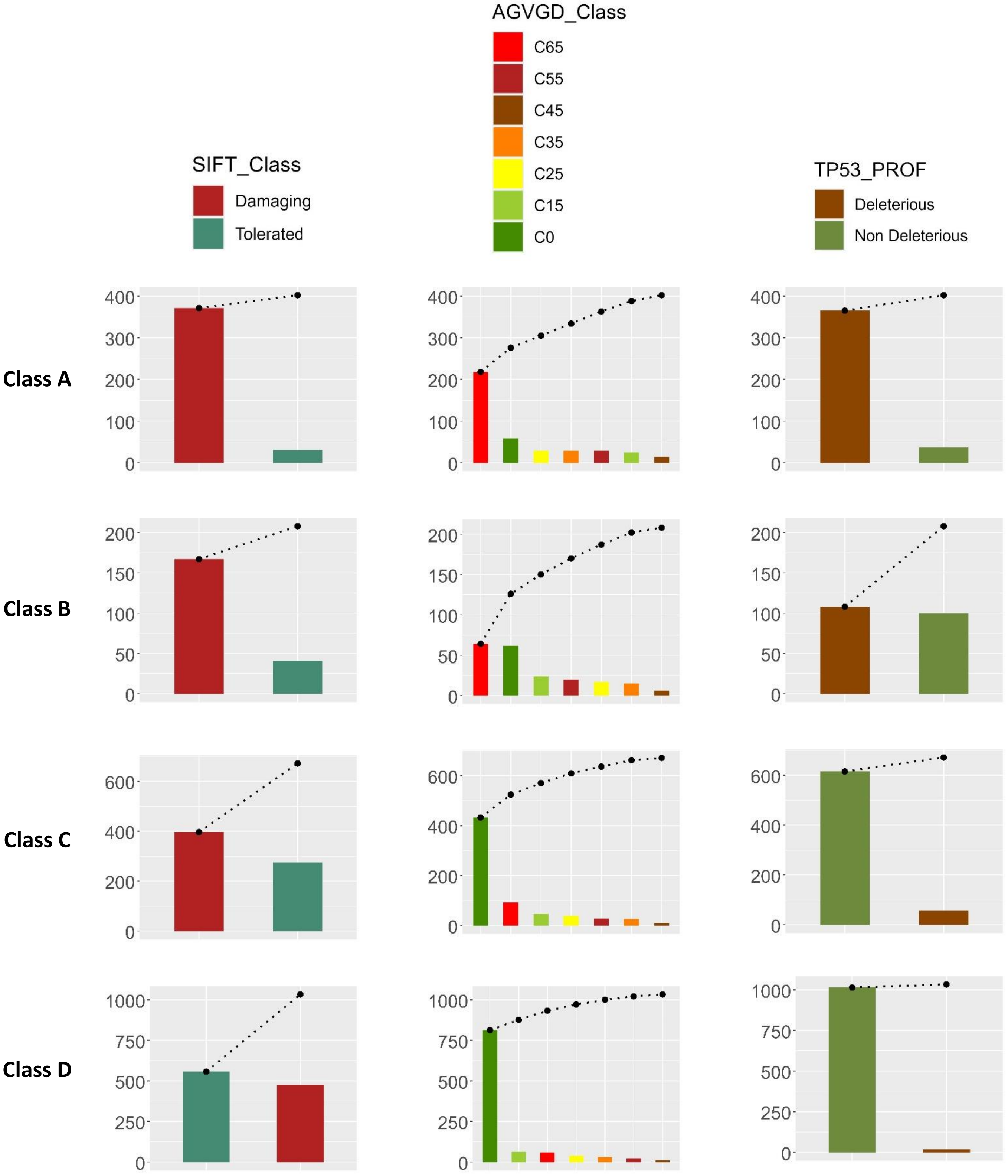

AGVGD_Class
C65
C55
C45
C35
C25
C15
C0
SIFT_Class
Damaging
Tolerated
TP53_PROF
Deleterious
Non Deleterious
Class A
Class B
Class C
Class D

**Figure S5**

**A.**

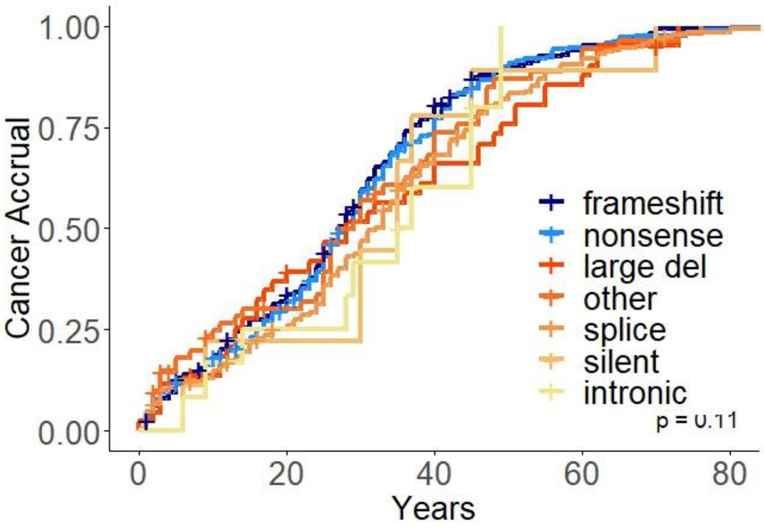


**B.**

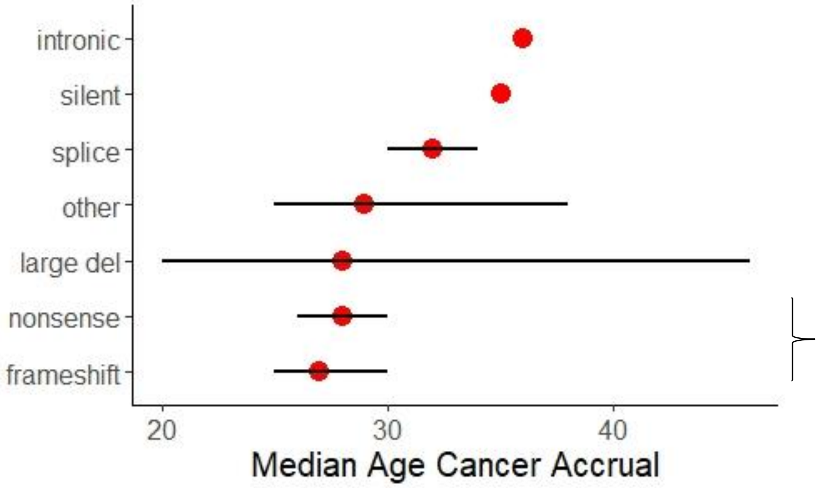


**Selection of null variants: nonsense and frameshift -> Class 0**

**Figure S6**

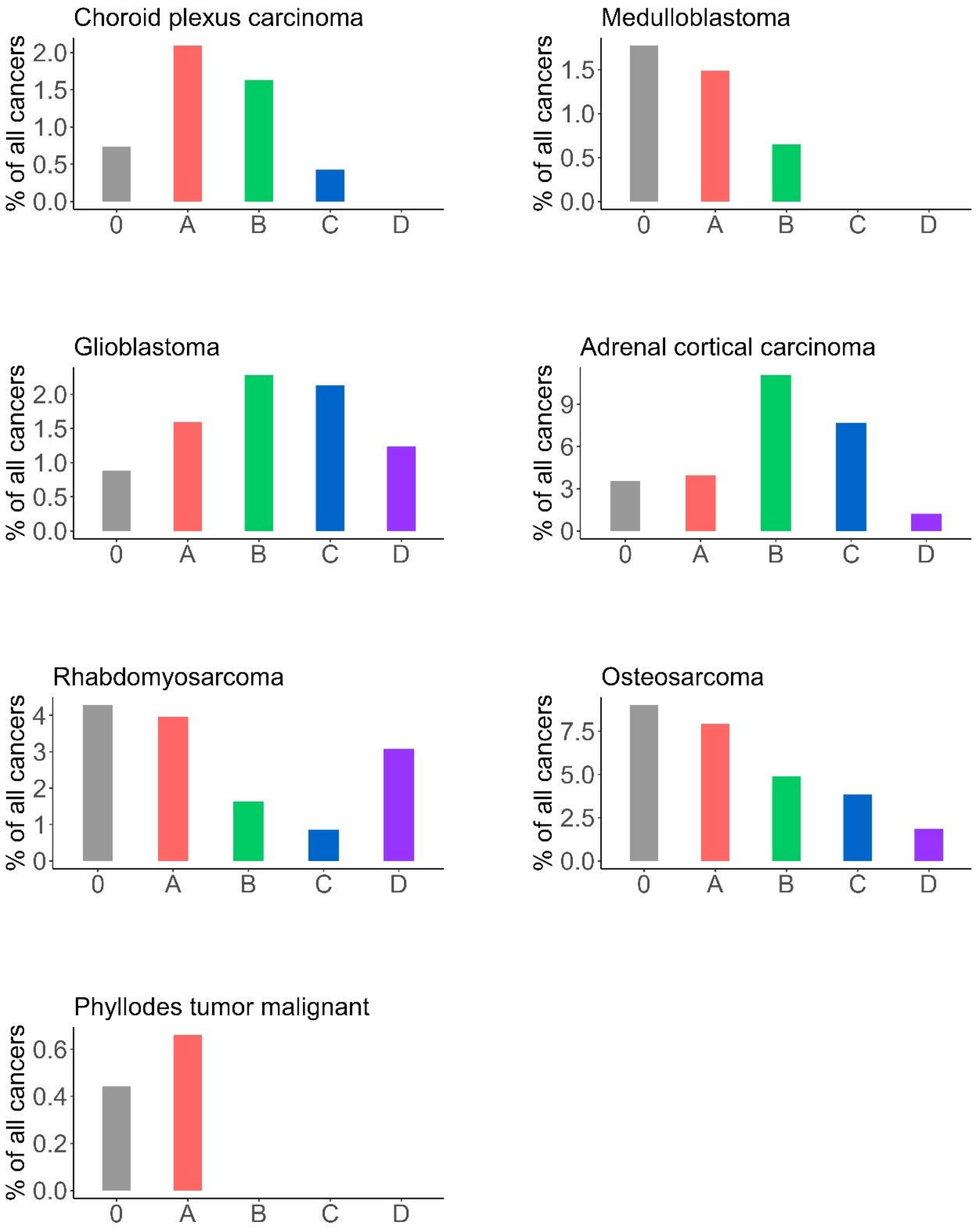

Choroid plexus carcinoma
% of all cancers
2.0
1.5
1.0
0.5
0.0
0
A
B
C
D
Medulloblastoma
% of all cancers
1.5
1.0
0.5
0.0
0
A
B
C
D
Glioblastoma
% of all cancers
2.0
1.5
1.0
0.5
0.0
0
A
B
C
D
Adrenal cortical carcinoma
% of all cancers
9
6
3
0
0
A
B
C
D
Rhabdomyosarcoma
% of all cancers
4
3
2
1
0
0
A
B
C
D
Osteosarcoma
% of all cancers
7.5
5.0
2.5
0.0
0
A
B
C
D
Phyllodes tumor malignant
% of all cancers
0.6
0.4
0.2
0.0
0
A
B
C
D

### Figure S7

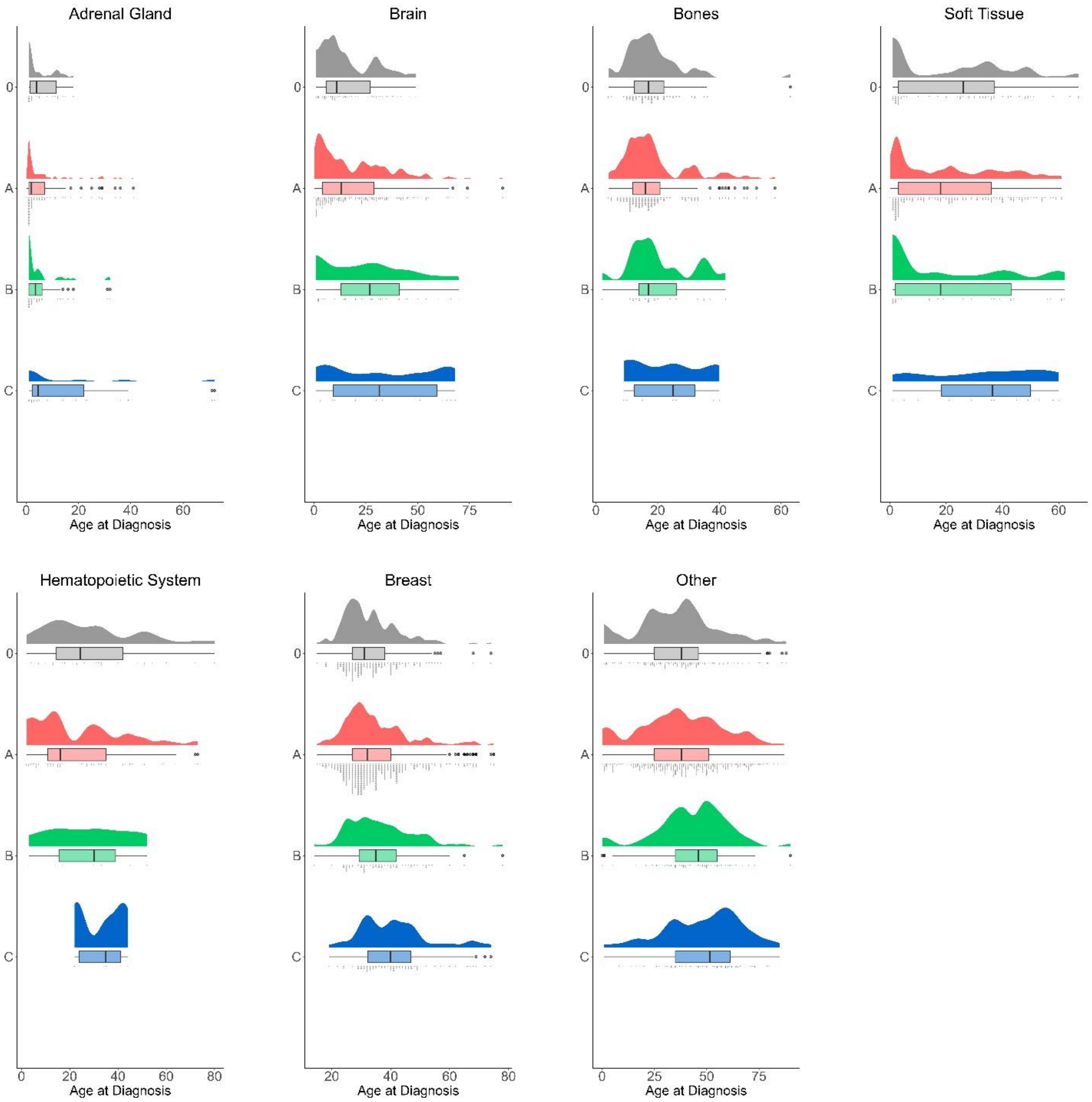
Adrenal Gland
Brain
Bones
Soft Tissue
Hematopoietic System
Breast
Other
0
A
B
C
Age at Diagnosis

### Figure S8

**A.**

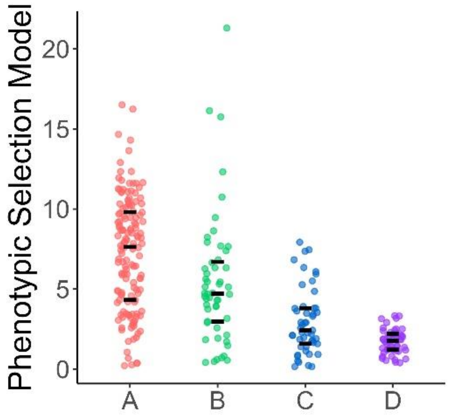


**Separation of classes in quartiles:**
**0-25% -> 1st quartile**
**25-50% -> 2nd quartile**
**50-75% -> 3rd quartile**
**75-100% -> 4th quartile**

**B.**

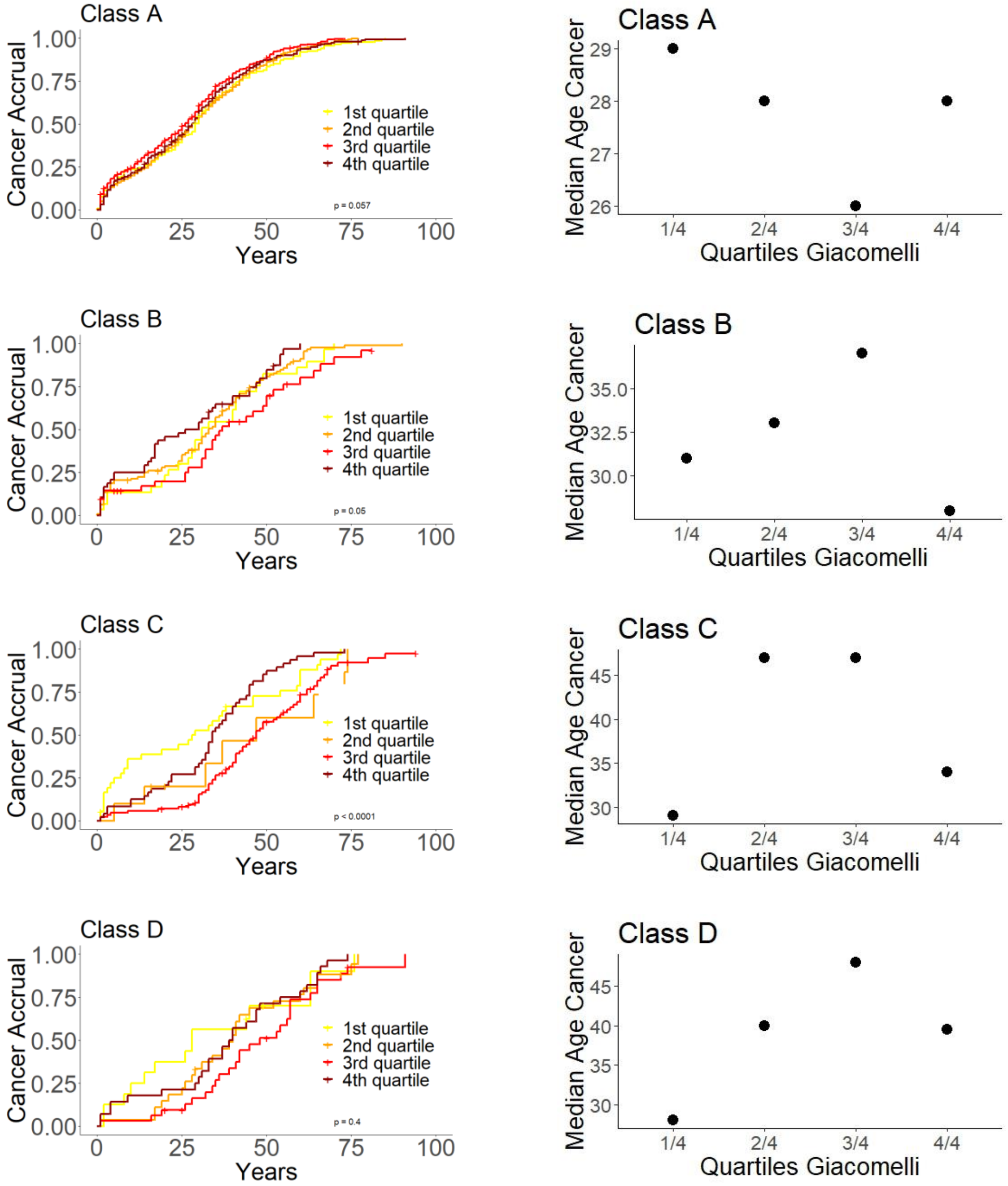

### Figure S9

**A.**

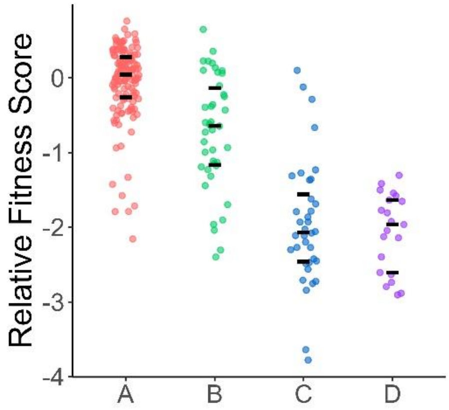


**Separation of classes in quartiles:**
**0-25% -> 1st quartile**
**25-50% -> 2nd quartile**
**50-75% -> 3rd quartile**
**75-100% -> 4th quartile**

**B.**

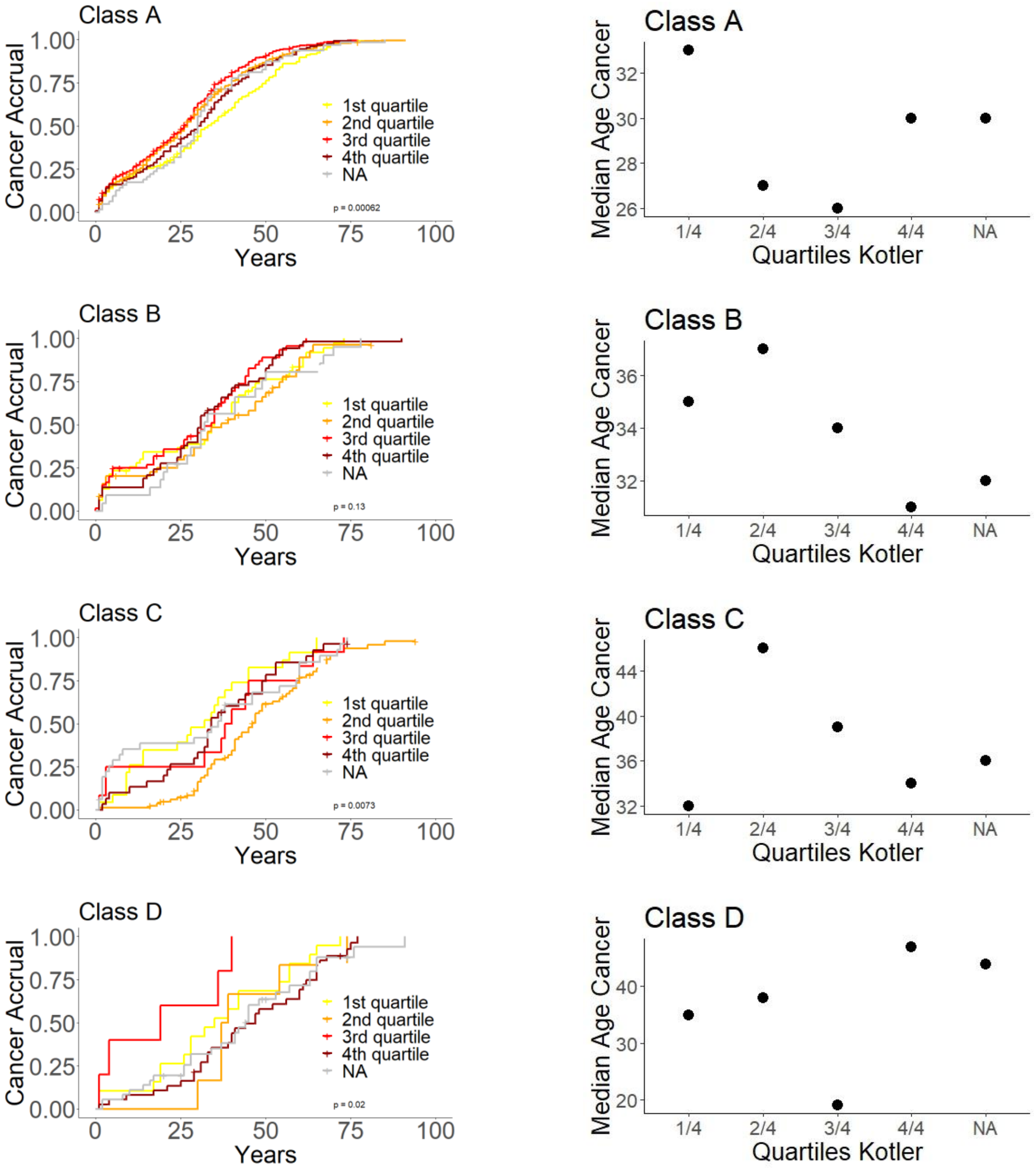

**Figure S10**

**A.**

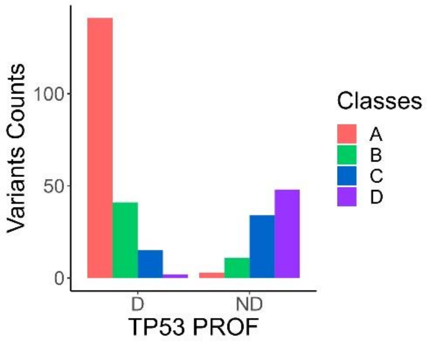


**B.**

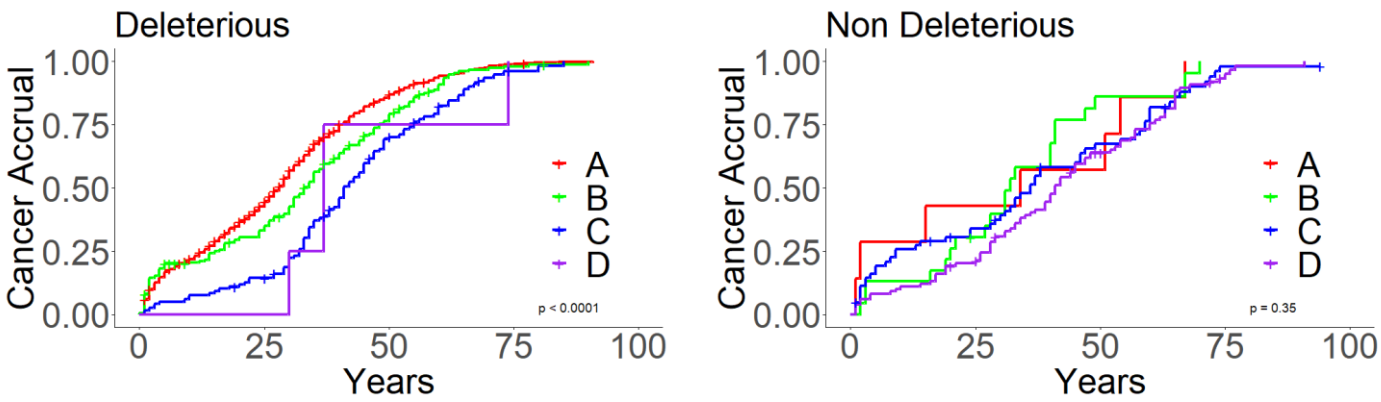

**Figure S11**

**A.** French Cohort

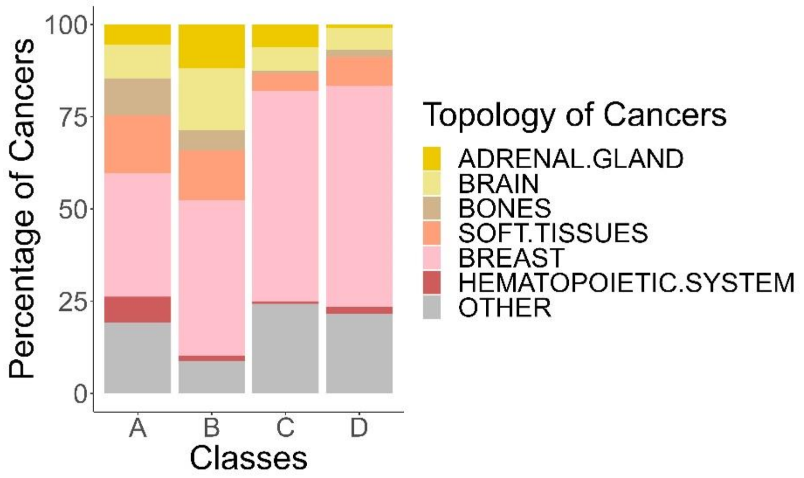


**B.** German Cohort

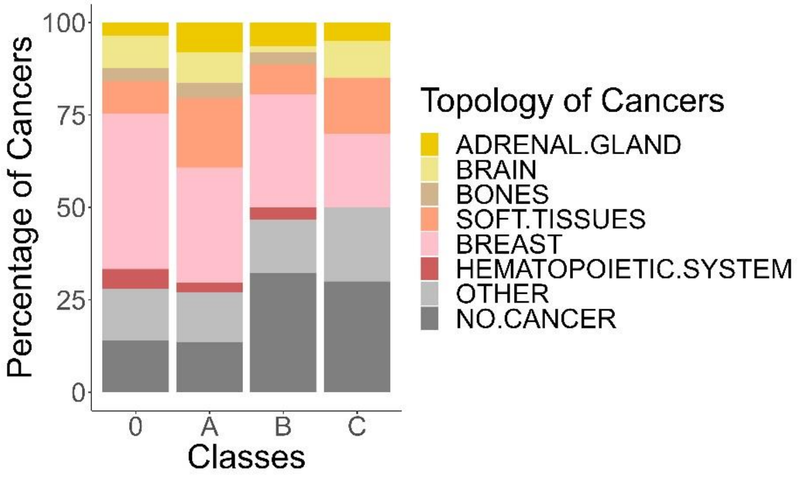


**C.** Canadian Cohort

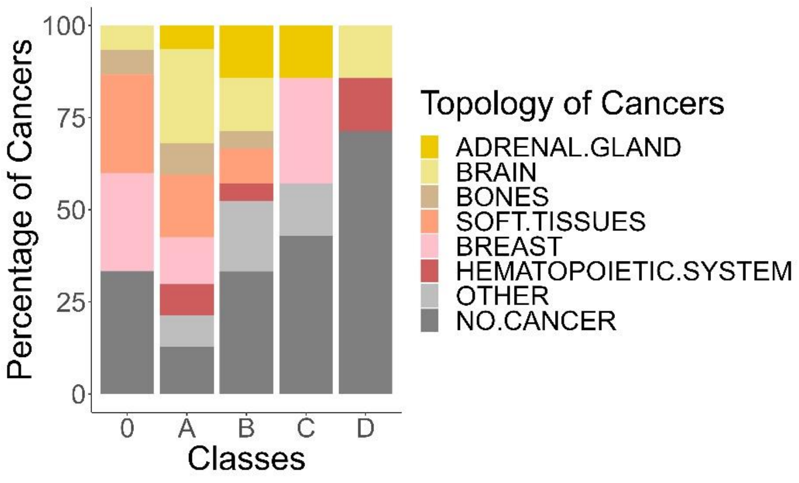
